# Fitting to the UK COVID-19 outbreak, short-term forecasts and estimating the reproductive number

**DOI:** 10.1101/2020.08.04.20163782

**Authors:** Matt J. Keeling, Louise Dyson, Glen Guyver-Fletcher, Alex Holmes, Malcolm G Semple, ISARIC4C Investigators, Michael J. Tildesley, Edward M. Hill

**Author notes:** These authors contributed equally to this work (Louise Dyson, Michael J. Tildesley, Edward M. Hill). Membership list can be found in the backmatter (ISARIC4C Investigators).

## Abstract

The COVID-19 pandemic has brought to the fore the need for policy makers to receive timely and ongoing scientific guidance in response to this recently emerged human infectious disease. Fitting mathematical models of infectious disease transmission to the available epidemiological data provides a key statistical tool for understanding the many quantities of interest that are not explicit in the underlying epidemiological data streams. Of these, the effective reproduction number, *R*, has taken on special significance in terms of the general understanding of whether the epidemic is under control (*R <* 1). Unfortunately, none of the epidemiological data streams are designed for modelling, hence assimilating information from multiple (often changing) sources of data is a major challenge that is particularly stark in novel disease outbreaks.

Here, focusing on the dynamics of the first-wave (March-June 2020), we present in some detail the inference scheme employed for calibrating the Warwick COVID-19 model to the available public health data streams, which span hospitalisations, critical care occupancy, mortality and serological testing. We then perform computational simulations, making use of the acquired parameter posterior distributions, to assess how the accuracy of short-term predictions varied over the timecourse of the outbreak. To conclude, we compare how refinements to data streams and model structure impact estimates of epidemiological measures, including the estimated growth rate and daily incidence.

## 1 Introduction

In late 2019, accounts emerged from Wuhan city in China of a virus of unknown origin that was leading to a cluster of pneumonia cases [1]. The virus was identified as a novel strain of coronavirus on 7th January 2020 [2], subsequently named Severe Acute Respiratory Syndrome Coronavirus (SARS-CoV-2), causing the respiratory syndrome known as COVID-19. The outbreak has since developed into a global pandemic. As of 3rd August 2020 the number of confirmed COVID-19 cases was approaching 18 million, with more than 685,000 deaths occurring worldwide [3]. Faced with these threats, there is a need for robust predictive models that can help policy makers by quantifying the impact of a range of potential responses. However, as is often stated, models are only as good as the data that underpins them; it is therefore important to examine, in some detail, the parameter inference methods and agreement between model predictions and data.

In the UK, the first cases of COVID-19 were reported on 31st January 2020 in the city of York. Cases continued to be reported sporadically throughout February and by the end of the month guidance was issued stating that travellers from the high-risk epidemic hotspots of Hubei province in China, Iran and South Korea should self-isolate upon arrival in the UK. By mid-March, as the number of cases began to rise, there was advice against all non-essential travel and, over the coming days, several social-distancing measures were introduced including the closing of schools, non-essential shops, pubs and restaurants. This culminated in the introduction of a UK lockdown, announced on the evening of 23rd March 2020, whereby the public were instructed to remain at home with four exceptions: shopping for essentials; any medical emergency; for one form of exercise per day; and to travel to work if absolutely necessary. By mid-April 2020, these stringent mitigation strategies began to have an effect, as the number of confirmed cases and deaths as a result of the disease began to decline. As the number of daily confirmed cases continued to decline during April, May and into June, measures to ease lockdown restrictions began, with the re-opening of some non-essential businesses and allowing small groups of individuals from different households to meet up outdoors, whilst maintaining social distancing. This was followed by gradually re-opening primary schools in England from 1st June 2020 and all non-essential retail outlets from 15th June 2020. Predictive models for the UK are therefore faced with a changing set of behaviours against which historic data must be judged, and an uncertain future of potential additional relaxations.

Throughout, a significant factor in the decision-making process was the value of the effective reproduction number, *R*, of the epidemic. The effective reproduction number is a time-varying measure of the average number of secondary cases per infectious case in a population (made up of both susceptible and non-susceptible hosts) and has been a quantity estimated by several modelling groups that provided advice through the Scientific Pandemic Influenza Modelling Group (SPI-M) [4]. Note, the effective reproduction number differs from the basic reproduction number, *R*_0_ (the average number of secondary infections produced by a typical case of an infection in a population where everyone is susceptible). The Warwick COVID-19 model presented here provided one source of *R* estimates through SPI-M. When *R* is estimated to be significantly below one, such that the epidemic is exponentially declining, then there is scope for some relaxation of intervention measures. However, as *R* approaches one, further relaxation of control may lead to cases starting to rise again. It is therefore crucial that models continue to be fitted to the latest epidemiological data in order for them to provide the most robust information regarding the impact of any relaxation policy and the effect upon the value of *R*. It is crucial to note, however, that there will necessarily be a delay between any change in behaviour, the epidemiological impact and the ability of a statistical method to detect this change.

The initial understanding of key epidemiological characteristics for a newly emergent infectious disease is, by its very nature of being novel, extremely limited and often biased towards early severe cases. Developing models of infectious disease dynamics enables us to challenge and improve our mechanistic understanding of the underlying epidemiological processes based on a variety of data sources. One way such insights can be garnered is through model fitting / parameter inference, the process of estimating the parameters of the mathematical model from data. The task of fitting a model to data is often challenging, partly due to the necessary complexity of the model in use, but also because of data limitations and the need to assimilate information from multiple sources of data [5].

Throughout this work, the process of model fitting is performed under a Bayesian paradigm, where knowledge of the parameters are modelled through random variables and have joint probability distributions [6]. In full, the posterior distribution of the parameters *θ* given the data, *P* (*θ*|*D*), describes how our prior beliefs in the distributional properties of the parameters, *P* (*θ*), have updated as a consequence of the information in the data, which is captured through the likelihood function (the probability distribution of the data given the model and parameters, *L*(*D*|*θ*))). Through applying Bayes’ theorem, the relationship between the posterior, the likelihood is encapsulated by,

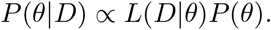

Whilst we would ideally seek an analytical expression for the target posterior distribution, in many cases the solution for the posterior distribution is not mathematically tractable. As a consequence, we revert to deriving empirical estimates of the desired probability distribution. In particular, we use Markov Chain Monte Carlo (MCMC) schemes to find the posterior probability distribution of our parameter set given the data and our prior beliefs. MCMC methods construct a Markov chain which converges to the desired posterior parameter distribution at steady state [7]. Simulating this Markov chain thus allows us to draw sets of parameters from the joint posterior distribution.

Adopting a Bayesian approach to parameter inference means parameter uncertainty may then be propagated if using the model to make projections. This affords models with mechanistic aspects, through computational simulation, the capability of providing an estimated range of predicted possibilities given the evidence presently available. Thus, models can demonstrate important principles about outbreaks [8], with examples during the present pandemic including analyses of the effect of non-pharmaceutical interventions on curbing the outbreak of COVID-19 in the UK [9].

In this paper, we present the inference scheme, and its subsequent refinements, employed for calibrating the Warwick SARS-CoV-2 transmission and COVID-19 disease model [10] to the available public health data streams and estimating key epidemiological quantities such as *R* during the first wave of SARS-CoV-2 infection in the UK (March-June 2020). In particular it is worth stressing that throughout we present our approach as it evolved during the outbreak, rather than the optimal methods and assumptions that would be made with hindsight. In addition, the paper was initially composed in July-August 2020 and we have largely retained the contextual information as originally written. In other words, we treat the manuscript as a record of the state of our modelling at that time.

We begin by describing our mechanistic transmission model for SARS-CoV-2 in Section 2, detailing in Section 3 how the effects of social distancing are incorporated within the model framework. In order to fit the model to data streams pertaining to critical care, such as hospital admissions and bed occupancy, Section 4 expresses how epidemiological outcomes were mapped onto these quantities. In Section 5, we outline how these components are incorporated into the likelihood function and the adopted MCMC scheme. The estimated parameters are then used to measure epidemiological measures of interest, such as the growth rate (*r*), with the approach detailed in Section 6.

The closing sections draw attention to how model frameworks may evolve during the course of a disease outbreak as more data streams become available and we collectively gain a better understanding of the epidemiology (Section 7). We explore how key epidemiological quantities, in particular the reproduction number *R* and the growth rate *r*, depend on the data sources used to underpin the dynamics (Section 8). To finish, we outline the latest fits and model generated estimates using data up to mid-June 2020 (Section 9).

## 2 Model description

Here we present the University of Warwick SEIR-type compartmental age-structured model, developed to simulate the spread of SARS-CoV-2 within regions of the UK. Matched to a variety of epidemiological data, the model operates and is fitted to data from the seven NHS regions in England (East of England, London, Midlands, North East and Yorkshire, North West, South East, South West) and the three devolved nations (Northern Ireland, Scotland and Wales). The model incorporates multiple layers of heterogeneity, through partitioning the population into five-year age classes, tracking symptomatic and asymptomatic transmission, accounting for household saturation of transmission and household quarantining.

The population is stratified into multiple compartments with respect to SARS-CoV-2 infection status (Fig. 1): individuals may be susceptible (*S*), exposed (*E*), with detectable infection (symptomatic *D*), or undetectable infection (asymptomatic, *U*). Undetectable infections are assumed to transmit infection at a reduced rate given by *τ*. We let superscripts denote the first infection in a household (*F*), a subsequent infection from a detectable/symptomatic household member (*SD*) and a subsequent infection from an asymptomatic household member (*SU*). A fraction (*H*) of the first detected case in a household is quarantined (*QF*), as are all their subsequent household infections (*QS*) - we ignore the impact of household quarantining on the susceptible population as the number in quarantine is assumed small compared with the rest of the population. The recovered class is not explicitly modelled, although it may become important once we have a better understanding of the duration of immunity. Natural demography and disease-induced mortality are ignored in the formulation of the epidemiological dynamics.

**Fig. 1:**
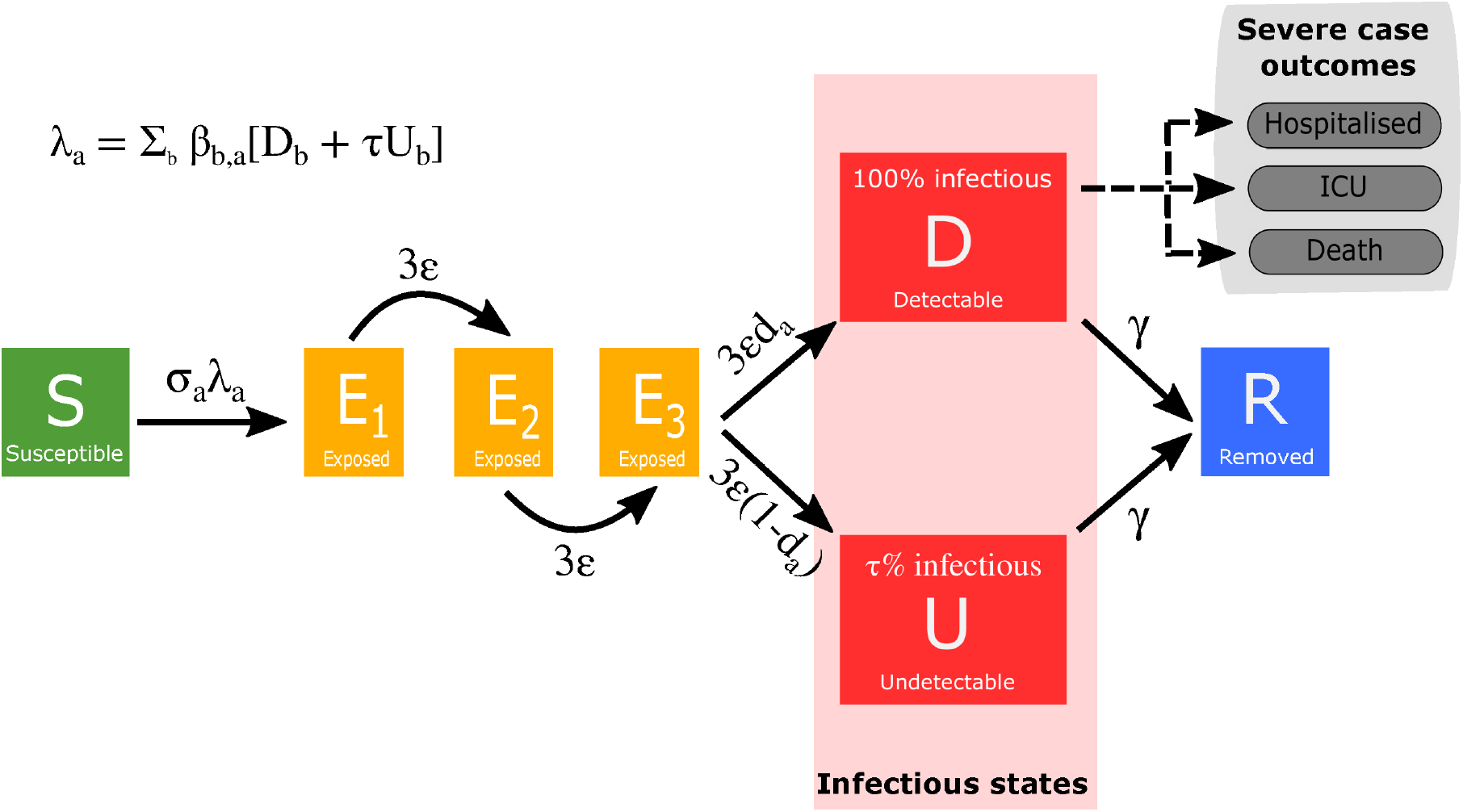
Model schematic of infection states and transitions. We stratified the population into susceptible, exposed, detectable infectious, undetectable infectious, and removed states. Solid lines correspond to disease state transitions, with dashed lines representing mapping from detectable cases to severe clinical cases that require hospital treatment, critical care (ICU), or result in death. We stratified the population into five year age brackets. See Tables 1 and 2 for a listing of model parameters. Note, we have not included quarantining or household infection status on this depiction of the system.

The model is deterministic in structure based on a large set of coupled ordinary differential equations (ODEs). Obviously, the continuous results from these ODEs are never going to precisely match the discrete integer-valued data, and hence we need a method of calculating the goodness of fit. We achieve this through a likelihood approach, assuming the data is Poisson distributed (or Binomial for quantities that have a relatively low upper bound) with a mean that is given by the results of the ODE model.

### Model equations

We provide a description of the model parameters in Tables 1 and 2. The full system of ordinary differential equations (ODEs) for the model are given by:

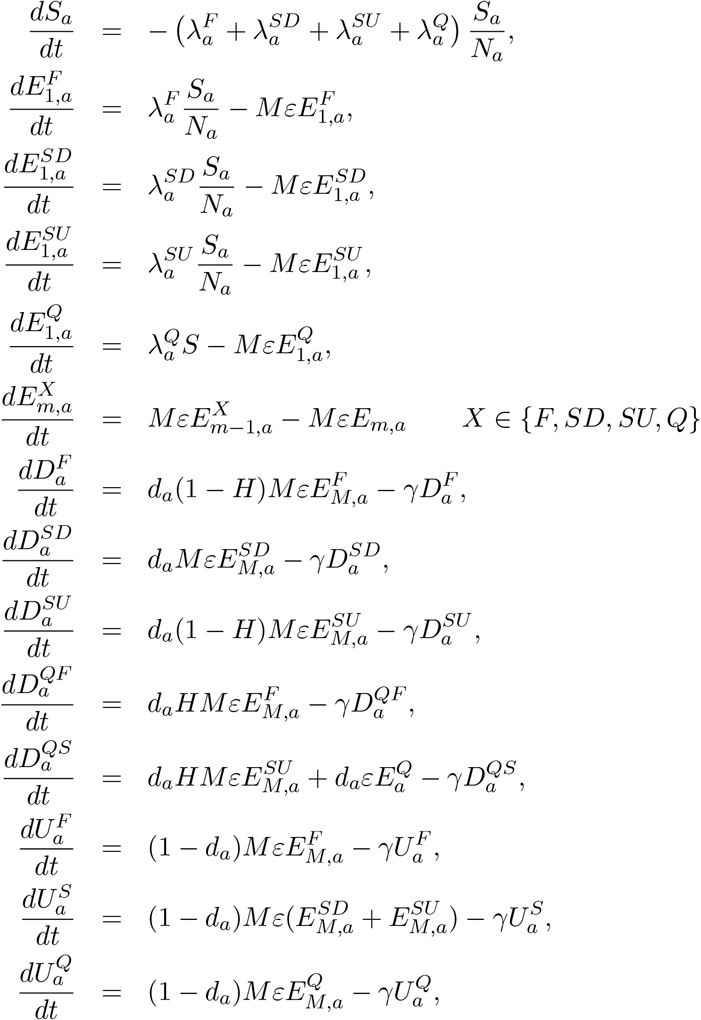

where *a* refers to each of the 21 5-year age groups (e.g. 0-4, 5-9 etc). We have included *M* latent classes for individuals infected with the virus but not yet infectious. The rate of progression from each latent class was *ϵM*, with the length of the total latent period being *ϵ*^−1^; in a stochastic framework this would be equivalent to the time in the latent class being an Erlang distribution with shape parameter *M* and rate parameter *ϵM*. Throughout we have taken *M* = 3. The rate of leaving the infectious class is *γ*; equivalent to an exponential distributed infectious period of length *γ*^−1^ in a stochastic framework.

**Table 1:** Description of key model parameters not fitted in the MCMC and their source

| Parameter | Description | Source |
| --- | --- | --- |
| $\beta$ | Age-dependent transmission, split into household, school, work and other | Matrices from Prem *et al.* [11] |
| $\gamma$ | Recovery rate, changes with $\tau$, the relative level of transmission from undetected asymptomatics compared to detected symptomatics | Fitted from early age-stratified UK case data to match growth rate and $R_0$ |
| $d_a$ | Age-dependent probability of displaying symptoms (and hence being detected), changes with $\alpha$ and $\tau$ | Fitted from early age-stratified UK case data to capture the age profile of infection. |
| $\sigma_a$ | Age-dependent susceptibility, changes with $\alpha$ and $\tau$ | Fitted from early age-stratified UK case data to capture the age profile of infection. |
| $H^R$ | Household quarantine proportion (set equal to $0.8\phi_t^R$) | Can be varied according to scenario |
| $N_a^R$ | Population size of a given age within each region | ONS |

**Table 2:**
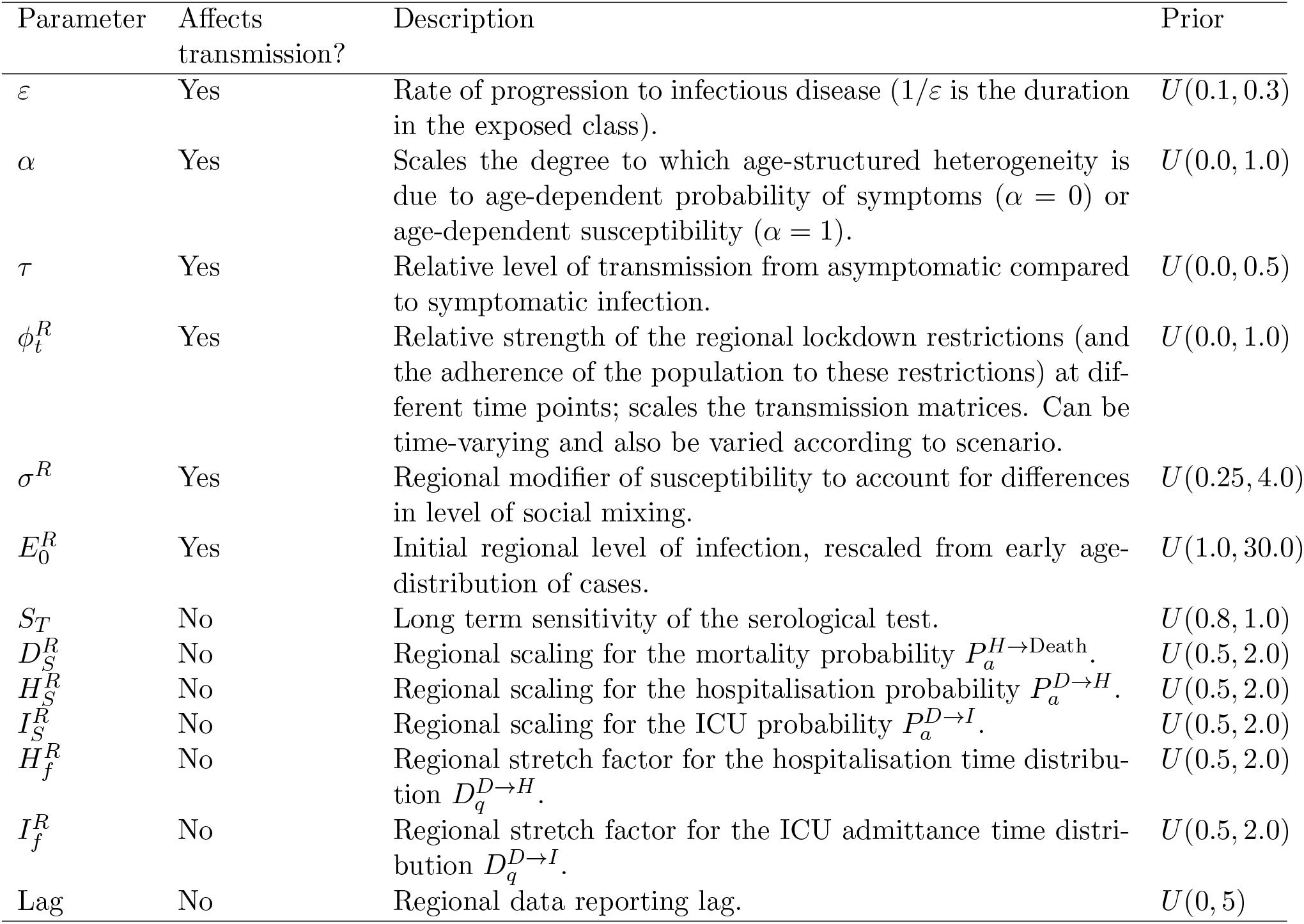
Description of key model parameters fitted in the MCMC

The forces of infection govern the non-linear transmission of infection. We partition the infectious pressure exerted on a given age group *a, λ*_*a*_, based on the category of infected case created: transmission in non-household settings generating first infecteds in households 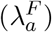, subsequent household infections caused by non-quarantined first infecteds in a household who are detectable/symptomatic 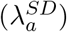, subsequent household infections caused by non-quarantined first infecteds in a household who are asymptomatic 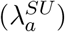, and subsequent household infections caused by quarantined first infecteds 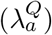. The collection of force of infection terms obey:

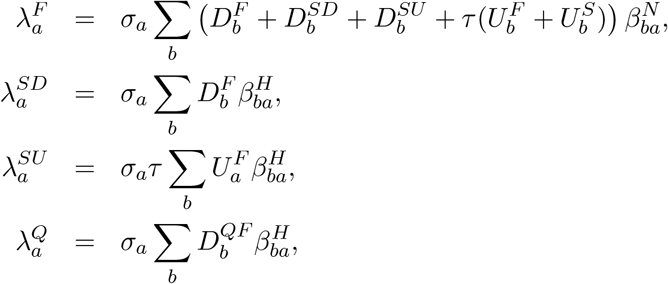

where 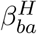 represents household transmission (with the subscript *ba* corresponding to transmission from age group *b* against age group *a*) and 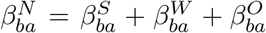 represents all other transmission locations, comprising school-based transmission 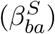, work-place transmission 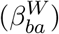 and transmission in all other locations 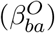. We took the setting specific age structured contact matrices from Prem et al [11], although other sources such as POLYMOD [12] could be used, with the modification of these contact patterns to model social distancing measures explained in Section 3. *σ*_*a*_ corresponds to the age-dependent susceptibility of individuals to infection, *d*_*a*_ the age-dependent probability of displaying symptoms (and hence being detected), and *τ* represents reduced transmission of infection by undetectable individuals compared to detectable infections.

### Amendments to within-household transmission

We wanted our model to be able to capture both individual level quarantining and isolation of households with identified cases. In a standard ODE framework the incorporation of household structure increases the dimensionality of the system. Combined with the inclusion of other heterogeneities, such as age structure, the result can be a system whose dimensionality results in model calibration and simulation only being achievable at large computational expense [13, 14]. Therefore, we instead make a number of approximations in our model to achieve a comparable effect.

We make the simplification that all within household transmission originates from the first infected individual within the household (denoted with a superscript *F* or *QF* if they quarantine). This allows us to assume that secondary infections within a household in isolation (denoted with a superscript *QS* or *Q*) play no further role in the transmission dynamics. This means that high levels of household isolation can drive the epidemic extinct, as only the first individual infected in each household can generate infections outside the household. This methodology also helps to capture to some degree household depletion of susceptibles (or saturation of infection), as secondary infections in the household are not able to generate additional household infections.

Given the novelty of the additional household structure that is included in this model, we clarify in more detail here the action of this formulation. We give a simpler set of equations (based on a standard *SIR* model) that contains a similar household structure; in particular, we take the standard *SIR* model and split the infected class into those first infected within a household (*I*_*F*_) and subsequent infections (*I*_*S*_):

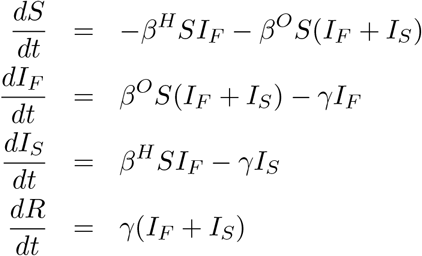

where the transmission rate is also split into within household transmission *β*^*H*^ and all other transmission *β*^*O*^ (i.e out-of-household transmission). Again, we make the assumption that only the first infection in any household generates infections within the household. We compare this to the SIR model without this additional structure:

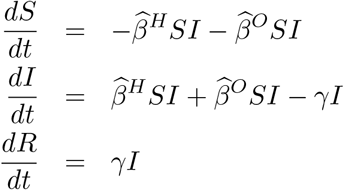

where we retain the split in transmission type.

The early growth rate of the two models are 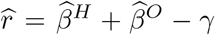for the simple SIR model, and 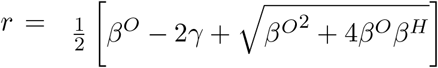 for the household structured version. From this simple comparison, it is clear that for the simple model the growth rate can remain positive even when control measures substantially reduce transmission outside the home (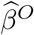 gets reduced), whereas in contrast for the structured version there is always a threshold level of transmission outside the household 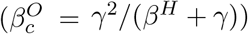 that is needed to maintain positive growth.

For both the simple household-structured model given here and the full COVID-19 model, the inclusion of additional household structure reduces the amount of within-household transmission compared to a model without household-structure — as only the initial infection in each household (*I*_*F*_) generates secondary within-household cases. It is therefore necessary to rescale the household transmission rate *β*^*H*^ to obtain the appropriate average within-household attack rate. For the full COVID-19 model, we found that a simple multiplicative scaling to the household transmission (*β*^*H*^ *→ zβ*^*H*^, *z* ≈ 1.3) generated a comparable match between the new model and a model without this household structure – even when age structure was included. We therefore included this scaling within the full model.

### Key Model Parameters

As with any model of this complexity, there are multiple parameters that determine the dynamics. Some of these are global parameters and apply for all geographical regions, with others used to capture the regional dynamics. Parameters that vary between regions are labelled with a superscript *R* defining the region of interest; other parameters are age-dependent, in which case we use subscript *a* to refer to the appropriate age-group. We separate two type of parameter that are required by our model formation. Those parameters in (Table 1) are generally from external sources and take fixed values (such as *β* or 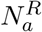), or are a fixed scaling of estimated values (such as *γ* or *H*^*R*^). In contrast a number of other parameters are inferred using the MCMC process (Table 2), some of which directly impact transmission and therefore determine the infection dynamics while others control the relationship between the infection dynamics and epidemiological observable quantities (such as the expected number of hospitalisations).

### Relationship between age-dependent susceptibility and detectability

We interlink age-dependent susceptibility, *σ*_*a*_, and detectability, *d*_*a*_, by a quantity *Q*_*a*_. *Q*_*a*_ can be viewed as the scaling between force of infection and symptomatic infection.

Further, in a population that may be divided into a finite number of discrete category according to a specific trait or traits (symptomatic and asymptomatic infection, for example), a next-generation approach can be used to relate the numbers of newly infected individuals in the various categories in consecutive generations [15]. Applying the next-generation approach to the symptomatic and asymptomatic infection states in our transmission model, the early dynamics would be specified by:

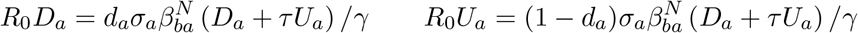

where *D*_*a*_ measures those with detectable infections, which mirrors the early recorded age distribution of symptomatic cases. Explicitly, we let 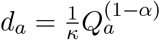 and 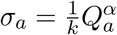. As a consequence, *Q*_*a*_ = *κkd* _*a*_ *σ*_*a*_ ; where the parameters *κ* and *k* are determined such that the oldest age groups have a 90% probability of being symptomatic (*d*_*>*90_ = 0.90) and such that the basic reproductive ratio from these calculations gives *R*_0_ = 2.7.

Throughout much of our work with this model, the values of *α* and *τ* are key in determining behaviour - in particular the role of school children in transmission [16]. We argue that a low *τ* and a low *α* are the only combination that are consistent with the growing body of data suggesting that levels of seroprevalence show only moderate variation across age-ranges [17], yet children do not appear to play a major role in transmission [18, 19]. To some extent, the separation into symptomatic (*D*) and asymptomatic (*U*) within the model is somewhat artificial as there are a wide spectrum of symptom severity that can be experienced.

### Regional Heterogeneity in the Dynamics

Throughout the current epidemic, there has been noticeable heterogeneity between the different regions of England and between the devolved nations. In particular, London is observed to have a large proportion of early cases and a relatively steeper decline in the subsequent lockdown than the other regions and the devolved nations. We capture this heterogeneity in our model through three estimated regional parameters that act on the heterogeneous population pyramid of each region.

Firstly, the initial level of infection in the region is re-scaled from the early age-distribution of cases, with the regional scaling factor 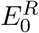 estimated by the MCMC process. Secondly, we allow the age-dependent susceptibility to be scaled between regions (scaling factor *σ*^*R*^) to account for different levels of social mixing and hence differences in the early *R*_0_ value. Finally, the relative strength of the lockdown (which may be time-varying) is again regional (scaling factor 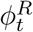) and also estimated by the MCMC process.

## 3 Modelling social distancing

We obtained age-structured contact matrices for the United Kingdom from Prem et al. [11], which we used to provide information on household transmission (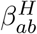, with the subscript *ab* corresponding to transmission from age group *a* against age group *b*), school-based transmission 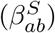, work-place transmission 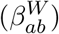 and transmission in all other locations 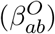.

We assumed that the suite of social-distancing and lockdown measures acted in concert to reduce the work, school and other matrices while increasing the strength of household contacts. Two additional parameters that acted to modulate the contact structure were the relative strength of lockdown interventions, *ϕ*_*t*_, and the proportion of work interactions that occur in public-facing ‘industries’, *θ* (we provide further details on both parameters later in this section).

We first capture the impact of social-distancing by defining new transmission matrices (*B*_*a,b*_), which represent the potential transmission in the presence of extreme lockdown. In particular, we assume that:

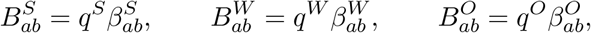

while household mixing *B*^*H*^ is increased by up to a quarter to account for the greater time spent at home. We set *q*^*S*^ = 0.05, *q*^*W*^ = 0.2 and *q*^*O*^ = 0.05 to approximate the reduction in attendance at school, attendance at workplaces and engagement with shopping and leisure activities in a maximum lockdown situation, respectively. Note that the parameterisation of the *q* parameters was subjective, with a higher value for the workplace setting used (corresponding to a lesser reduction in contacts) compared to all other settings on the basis of essential businesses maintaining a semblance of in-person staff attendance.

We used the assumed transmission matrices for a maximum lockdown scenario (*B*_*a,b*_) to generate new transmission matrices in each setting 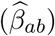 for a given strength of interventions and adherence level, *ϕ*_*t*_, as follows:

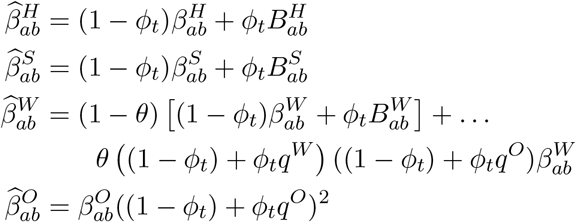

As such, home and school interactions are scaled between their pre-lockdown values (*β*) and post-lockdown limits (*B*) by the intervention and adherence parameter *ϕ*_*t*_. Work interactions that are not in public-facing ‘industries’ (a proportion 1 - *θ*) were also assumed to scale in this manner; while those that interact with the general populations (such as shop-workers) were assumed to scale as both a function of their reduction and the reduction of others. We have assumed *θ* = 0.3 throughout, which we subjectively chose, with us acknowledging that the use of an alternative parameterisation could alter the outcomes. Similarly, the reduction in transmission in other settings (generally shopping and leisure) has been assumed to scale with the reduction in activity of both members of any interaction, giving rise to a squared term.

## 4 Public Health Measurable Quantities

The main model equations focus on the epidemiological dynamics, allowing us to compute the number of symptomatic and asymptomatic infectious individuals over time. However, these quantities are not measured - and even the number of confirmed cases (the closest measure to symptomatic infections) is highly biased by the testing protocols at any given point in time. It is therefore necessary to convert infection estimates into quantities of interest that can be compared to data. We considered six such quantities which we calculated from the number of newly detectable symptomatic infections on a given day *nD*_*d*_.

### 1. Hospital Admissions

We assume that a fraction 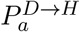 of detectable cases will be admitted into hospital after a delay *q* from the onset of symptoms. The delay, *q*, is drawn from a distribution 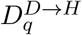 (note that 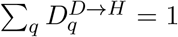.) Hospital admissions on day *d* of age *a* are therefore given by

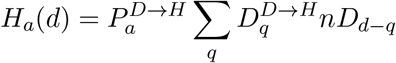

### 2. ICU Admissions

Similarly, a fraction 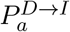 of detectable cases will be admitted into ICU after a delay, drawn from a distribution 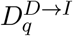 which determines the time between the onset of symptoms and admission to ICU. ICU admissions on day *d* of age *a* are therefore given by

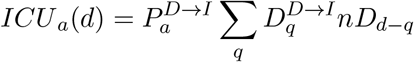

### 3. Hospital Beds Occupied

Individuals admitted to hospital spend a variable number of days in hospital. We therefore define two weightings, which determine if someone admitted to hospital still occupies a hospital bed *q* days later 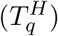 and if someone admitted to ICU occupies a hospital bed on a normal ward *q* days later 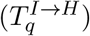. Hospital beds occupied on day *d* of age *a* are therefore given by

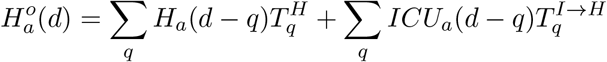

### 4. ICU Beds Occupied

We similarly define 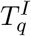as the probability that someone admitted to ICU is still occupying a bed in ICU *q* days later. ICU beds occupied on day *d* of age *a* are therefore given by

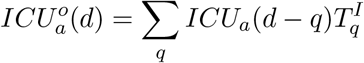

### 5. Number of Deaths

The mortality ratio 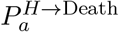 determines the probability that a hospitalised case of a given age, *a*, dies after a delay, *q* drawn from a distribution, 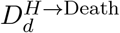 between hospitalisation and death. The number of deaths on day *d* of age *a* are therefore given by

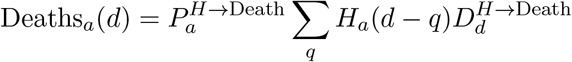

We note that while in the early stages of the epidemic only deaths from hospitalised individuals were initially registered as a death due to COVID, here we use all COVID deaths irrespective of where they occur. This measure has since been superseded by deaths within 28 days of a positive COVID test as a standardised measure in the UK. Therefore, 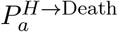 should be viewed as a relative scaling rather than an absolute probability that a hospitalised individual dies.

### 6. Proportion testing seropositive

Seropositivity is a function of time since the onset of symptoms; we therefore define an increasing sigmoidal function which determines the probability that someone who first displayed symptoms *q* days ago would generate a positive serology test from a blood sample. We matched the shape of this sigmoidal function to data from PHE (estimated independently, not within our MCMC scheme), while the asymptote (the long-term sensitivity of the test, *S*_*T*_) is a free parameter determined by the MCMC. We match our age-dependent prediction against antibody seroprevalence from weekly blood donor samples from different regions of England (approximately 1000 samples per region) [20].

These nine distributions are all parameterised from individual patient data as recorded by the COVID-19 Hospitalisation in England Surveillance System (CHESS) [21], the ISARIC WHO Clinical Characterisation Protocol UK (CCP-UK) database sourced from the COVID-19 Clinical Information Network (CO-CIN) [22, 23], and the PHE sero-surveillance of blood donors [20]. CHESS data is used to define the probabilities of different outcomes 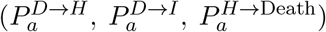 due to its greater number of records, while CCP-UK is used to generate the distribution of times 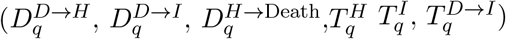 due to its greater detail (Fig. S1).

However, these distributions all represent a national average and do not therefore reflect regional differences. We therefore define regional scalings of the three key probabilities (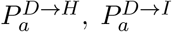 and 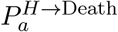) and two additional parameters that can stretch (or contract) the distribution of times spent in hospital and ICU. We infer these five regional parameters (Table 2), which are necessary to get good agreement between key observations in all regions and may reflect both differences in risk groups (in addition to age) between regions or differences in how the data are recorded between devolved nations. We stress that these parameters do not (of themselves) influence the epidemiological dynamics, but do strongly influence how we fit to the evolving dynamics.

## 5 Likelihood Function and the MCMC process

Multiple components form the likelihood function; most of which are based on a Poisson-likelihood. For brevity we define *L*_*P*_ (*n*|*x*) = (*n* ln(*x*) − *x*) − log(*n*!) as the log of the probability of observing *i* given a Poisson distribution with mean *x*. Similarly *L*_*B*_(*n*|*N, p*) = *nlog*(*p*) + (*N* − *n*)*log*(1 − *p*) is the log of the binomial probability function. The log-likelihood function is then:

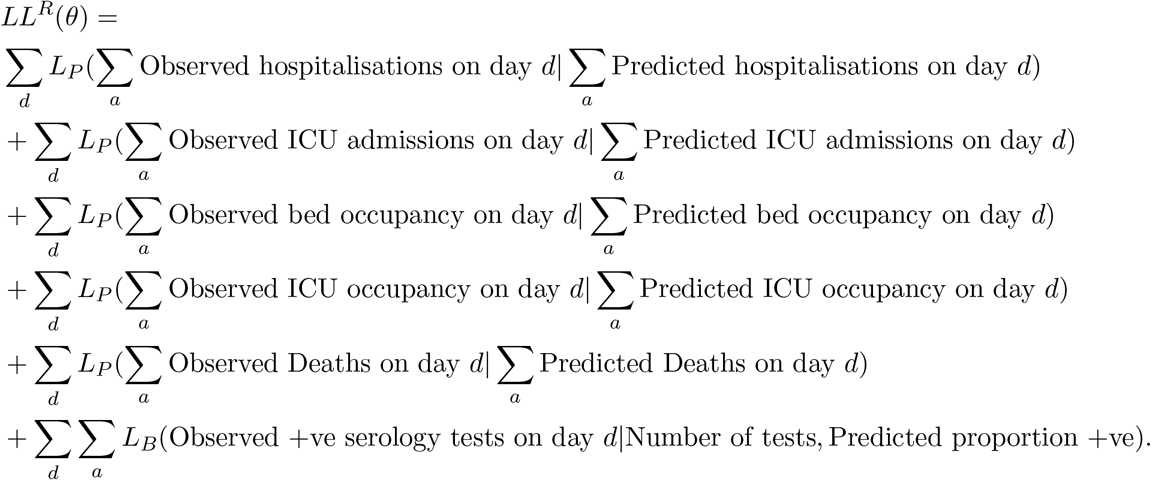

This log-likelihood is the key component of the MCMC scheme. In the MCMC process, we apply multiple updates of the parameters using normal or log-normal proposal distributions about the current values. Some parameters (the scaling of age-structure *α*, the relative transmission rate *τ*, the latent period 1*/ε* and the test sensitivity *S*_*T*_) are global and apply to all regions; new values of these are proposed and the log-likelihood calculated over all 10 regions. Other parameters are regional (such as the relative strength of lockdown restrictions 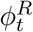) and can be updated for each region in turn, the ODEs simulated and stored. Finally, another set of regional parameters govern how the ODE output is translated into public health measurable quantities (Section 4). These can be rapidly applied to the solution to the ODEs and the likelihood calculated. Given the speed of this last set, multiple proposals are tested for each ODE replicate. We remark that the observation processes for the different public health measurable quantities are conditionally independent given the mechanistic model predictions.

New data are available on a daily time-scale, and therefore inference needs to be repeated on a similar time-scale. We can take advantage of this sequential refitting, taking random draws from the posteriors of the previous inference process to set the initial conditions for each chain, thus reducing the need for a long burn-in period.

## 6 Measuring the Growth Rate, *r*

The growth rate, *r*, is defined as the rate of exponential growth (*r >* 0) or decay (*r <* 0); and can be visualised as the gradient when plotting observables on a logarithmic scale. Figure 2 shows a simple example, whereby linear trends are fitted to the number of daily hospital admissions (per 100,000 people) in London. In this figure, three trend lines are plotted: one before lock-down; one during intense lock-down; and one after partial relaxation on 11th May 2020. This plot clearly highlights the very different speeds between the initial rise and the long-term decline.

**Fig. 2:**
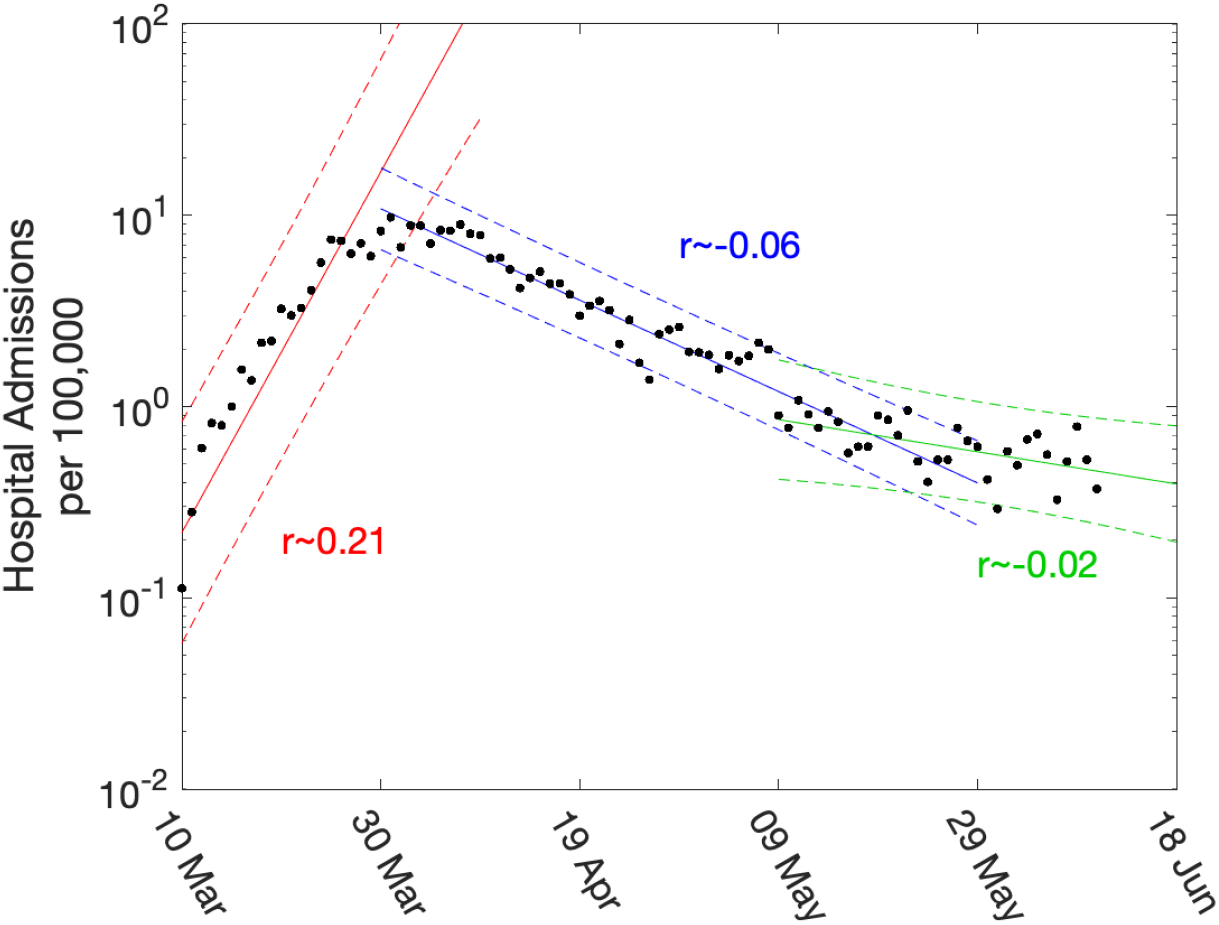
Daily hospital admissions per 100,000 individuals in London. Points show the number of daily admissions to hospital (both in-patients testing positive and patients entering hospital following a positive test); results are plotted on a log scale. We show three simple fits to the data are shown for pre-lockdown (red), strict-lockdown (blue) and relaxed-lockdown phases (green). Lines are linear fits to the logged data together with 95% confidence intervals, returning average growth rates of 0.21 (doubling every 3.4 days), −0.06 (halving every 11.5 days) and −0.02 (halving every 34 days).

While such statistically simple approaches are intuitively appealing, there are three main drawbacks. Firstly, they are not easily able to cope with the distributed delay between a change in policy (such as the introduction of the lockdown) and the impact of observable quantities (with the delay to deaths being multiple weeks). Secondly, they cannot readily utilise multiple data streams. Finally, they can only be used to extrapolate into the future - extending the period of exponential behaviour - they cannot predict the impact of further changes to policy. Our approach is to instead fit the ODE model to multiple data streams, and then use the daily incidence to calculate the growth rate. Since we use a deterministic set of ODEs, the instantaneous growth rate *r* can be calculated on a daily basis.

There has been a strong emphasis (especially in the UK) on the value of the reproductive number (*R*) which measures the expected number of secondary cases from an infectious individual in an evolving outbreak. *R* brings together both the observed epidemic dynamics and the time-frame of the infection, and is thus subject to uncertainties in the latent and infectious periods as well as in their distribution - although the growth rate and the reproductive number have to agree at the point *r* = 0 and *R* = 1. We have two separate methods for calculating *R* which have been found to be in very close numerical agreement. The first is to calculate *R* from the next generation matrix *β*_*ba*_*/γ* using the current distribution of infection across age-classes and states. The second (and numerically simpler method) is to use the relationship between *R* and *r* for an SEIR-type model with multiple latent classes, which gives

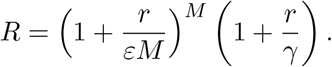

## 7 An Evolving Model Framework

Unsurprisingly, the model framework has evolved during the epidemic as more data streams have become available and as we have gained a better understanding of the epidemiology. Early models were largely based on the data from Wuhan, and made relatively crude assumptions about the times from symptoms to hospitalisation and death. Later models incorporated more regional variation, while the PHE serology data in early May 2020 had a profound impact on model parameters.

Figure 3 shows how our short-term predictions (each of three-weeks duration) changed over time, focusing on hospital admissions in London. It is clear that the early predictions were pessimistic about the reduction that would be generated by lockdown, although in part the higher values from early predictions is due to having identical parameters across all regions in the earliest models. In general later predictions, especially after the peak, are in far better agreement although the early inclusion of a step-change in the strength of the lockdown restrictions from 13th May 2020 (orange) led to substantial over-estimation of future hospital admissions. Across all regions we found some anomalous fits, which are due to changes in the way data were reported (Figs. S3 and S4).

**Fig. 3:**
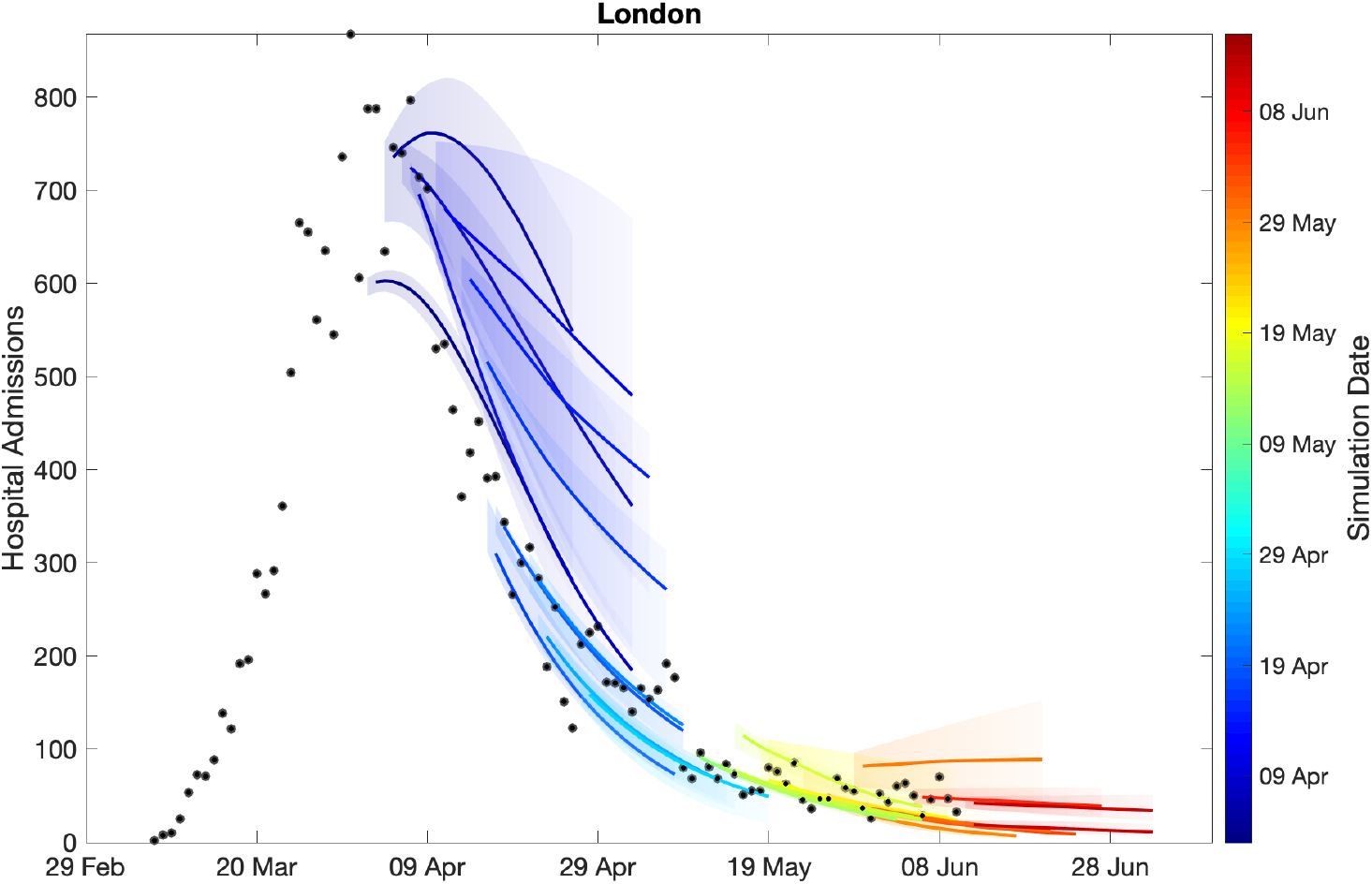
Sequential comparison of model results and data. For all daily hospital admissions with COVID-19 in London, we show the raw data (black dots) and a set of short-term predictions generated at different points during the outbreak. Changes to model fit reflect both improvements in model structure as well as increased amounts of data. The intervals represent our confidence in the fitted ODE model, and do not account for either stochastic dynamics nor the observational distribution about the deterministic predictions - which would generate far wider intervals.

The comparison of models and data over time can be made more formal by considering the mean squared error across the three-week prediction period for each region (Fig. 4). We compare three time varying quantities: (i) the mean value of the public health observable (in this case hospital deaths) in each region; (ii) the mean error between this data and the posterior set of ODE model predictions predicting forwards for three weeks; (iii) the mean error between the data and a simple moving average across the three time points before and after the data point. In each panel, the solid line corresponds to where the presented error statistic is equal to the mean, which is to be expected if the error originates from a Poisson distribution. The top left hand graph in Fig. 4 shows a clear linear relationship and correspondence in magnitude between the mean value and the error from the moving average, implying similarity between the variance and mean, giving support to our assumption (in the likelihood function) that the data are reasonably approximated as Poisson distributed. The other two graphs show how the error in the prediction has dropped over time from very high values for simulations in early April 2020 (when the impact of the lockdown was uncertain) to values in late May and June 2020 that are comparable with the error from the moving average.

**Fig. 4:**
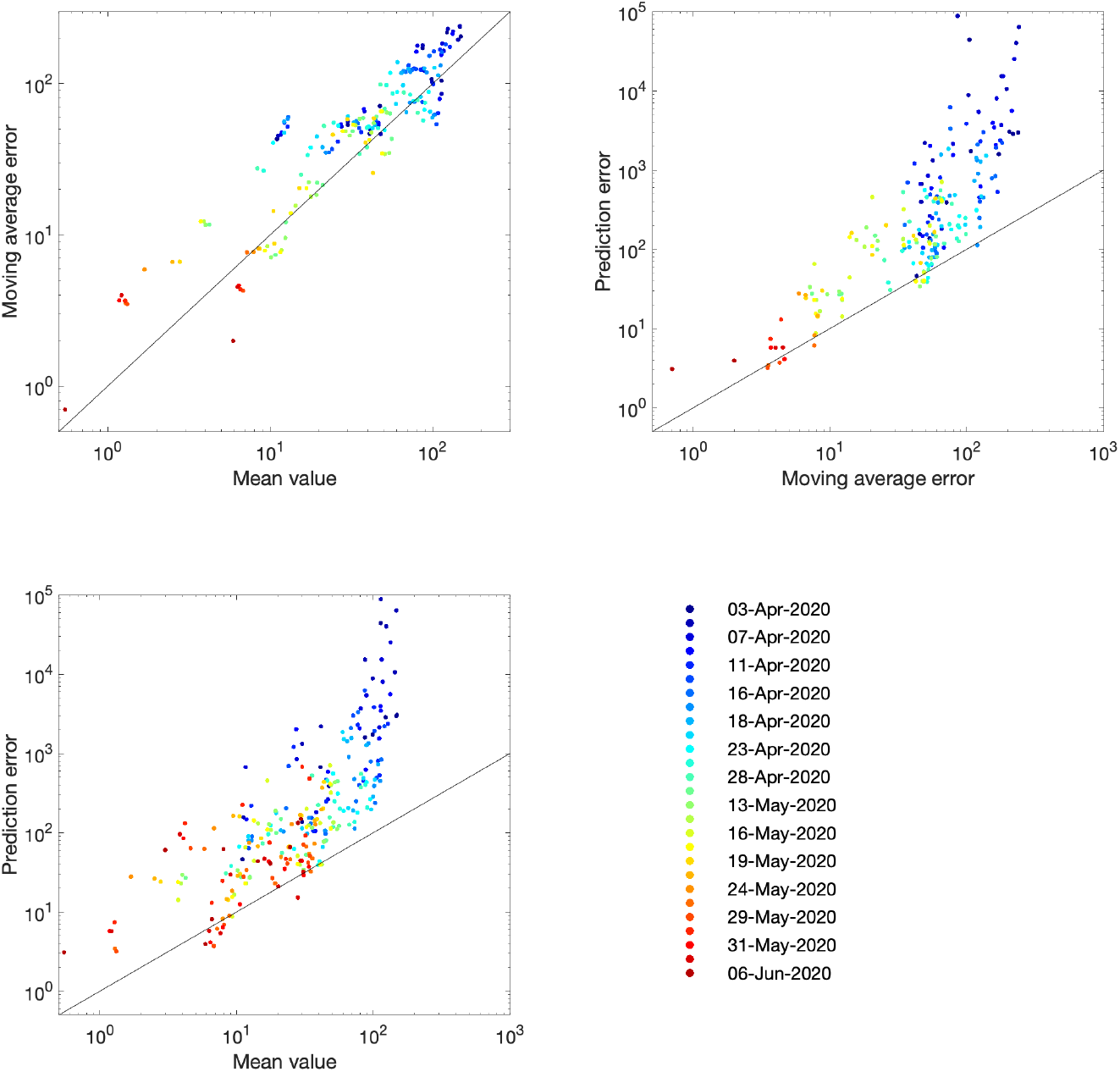
Improvement in fit over time for the number of hospital deaths. Each dot represents an analysis date (colour-coded) and region. For a data stream *x*_*t*_ and model replicates 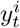 (where *i* accounts for sampling across the posterior parameter values) we compute the mean 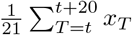; the prediction error 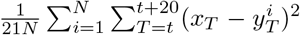; the moving average 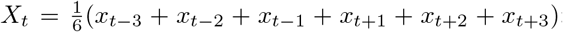; and the moving average error 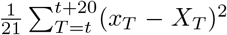. In each panel, the solid line corresponds to the path where the presented error statistic is equal to the mean.

## 8 Choice of Data Streams to Inform the Likelihood

The likelihood expression given above is an idealised measure, and depends on all the observed data streams being available and unbiased. Unfortunately, ICU admission data had not been available and there were subtle differences in data streams between the devolved nations. An important question is therefore how key epidemiological quantities (and in particular the reproduction number *R* and the growth rate *r*) depend on the data sources used to underpin the dynamics.

In a high dimensional systems with different temporal lags (see Fig. S1), there are inevitably different time-scales from when a change in policy or adherence occurs and when its impact is observed in key quantities. We briefly assess this problem in Figure 5, by considering the model output as surrogate data and examining how long a change in policy would take to impact the growth rate of key quantities. At time *t* = 0 we introduce a step-change in the strength of lockdown restrictions (*ϕ*_*t*_) within the model and recording the subsequent growth rates (*r*) associated with five key model outputs (infections, symptomatic cases, hospitalisations, admission to ICU and Deaths). Unsurprisingly, the impact of this change in restrictions takes the longest time to resolve in the mortality, taking around seven weeks for the estimate of the growth rate to stabilise to the asymptotic value. Even measures which should be more immediate, such as the growth of symptomatic cases, take some time to settle to the theoretical value (or *r* ≈ 0.01) given the high dimensionality of the age-structured model. This all strongly suggests that at best our estimates of *r* and hence *R* may not be able to rapidly detect changes to the underlying behaviour.

**Fig. 5:**
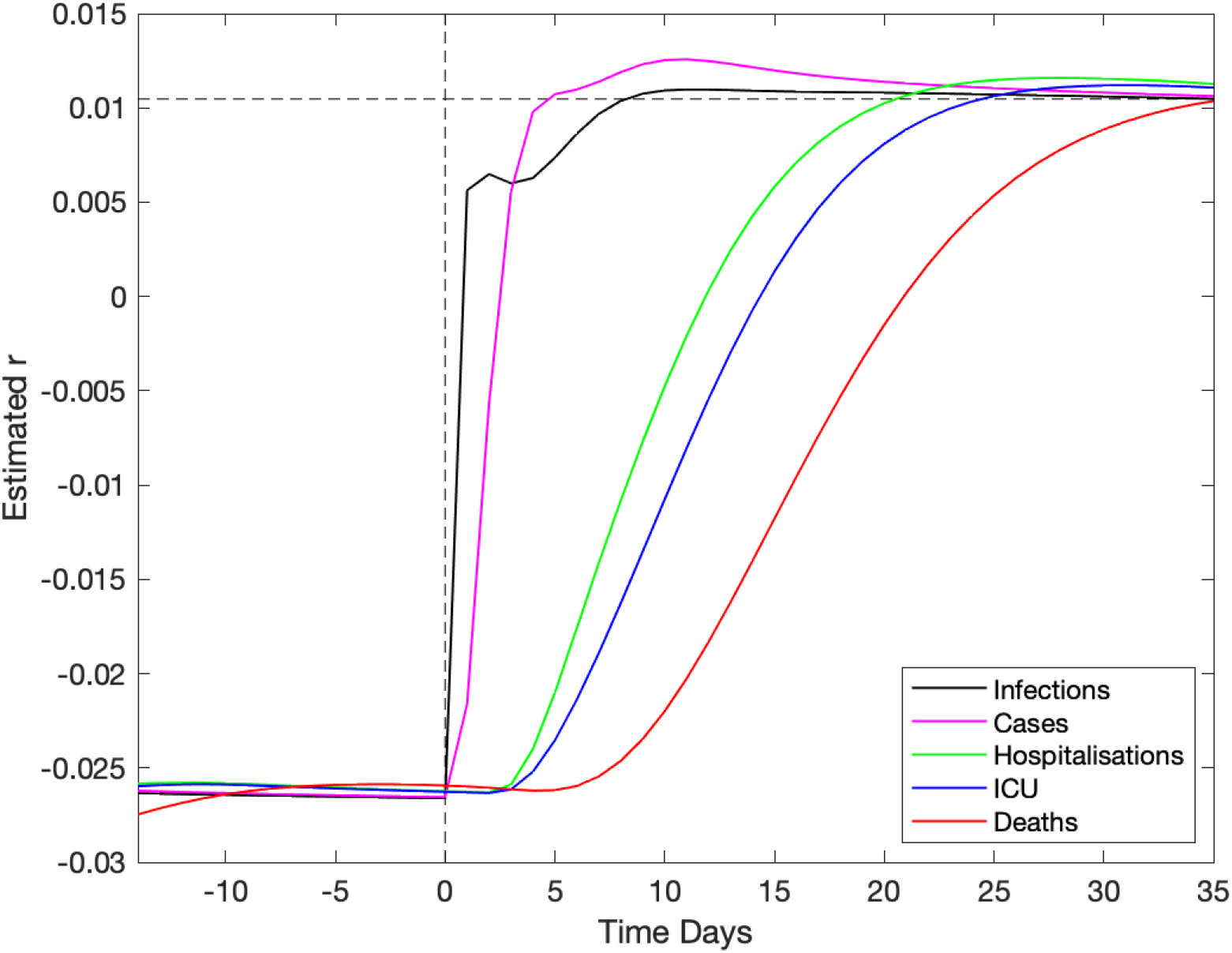
Impact of a change in underlying restrictions on the growth rate of modelled data streams. A change in the underlying restrictions occurs at time zero, taking the asymptotic growth rate *r* from ≈ − 0.026 to ≈+0.01. This change is reflected in an increase in growth rate of five key epidemiological quantities, which reach the true theoretical growth rate at different times.

Figure 6 (left panel) shows the impact of using different observables for London (other regions are shown in Fig. S5). This is achieved by only retaining a limited number of elements in the log-likelihood function, such that the model is matched to different combinations of data streams. Five different choices are shown: matching to recorded deaths only (using the date of death); matching to hospital admissions (both in-patients testing positive and admissions of individuals who have already tested positive); matching to bed occupancy, both hospital wards and ICU; matching to a combination of deaths and admissions; and finally matching to all data. Each of these different likelihood functions required an independent set of MCMC chains to be generated; from the associated posteriors we consider an estimate of the instantaneous growth rate as the most important epidemiological characteristic.

**Fig. 6:**
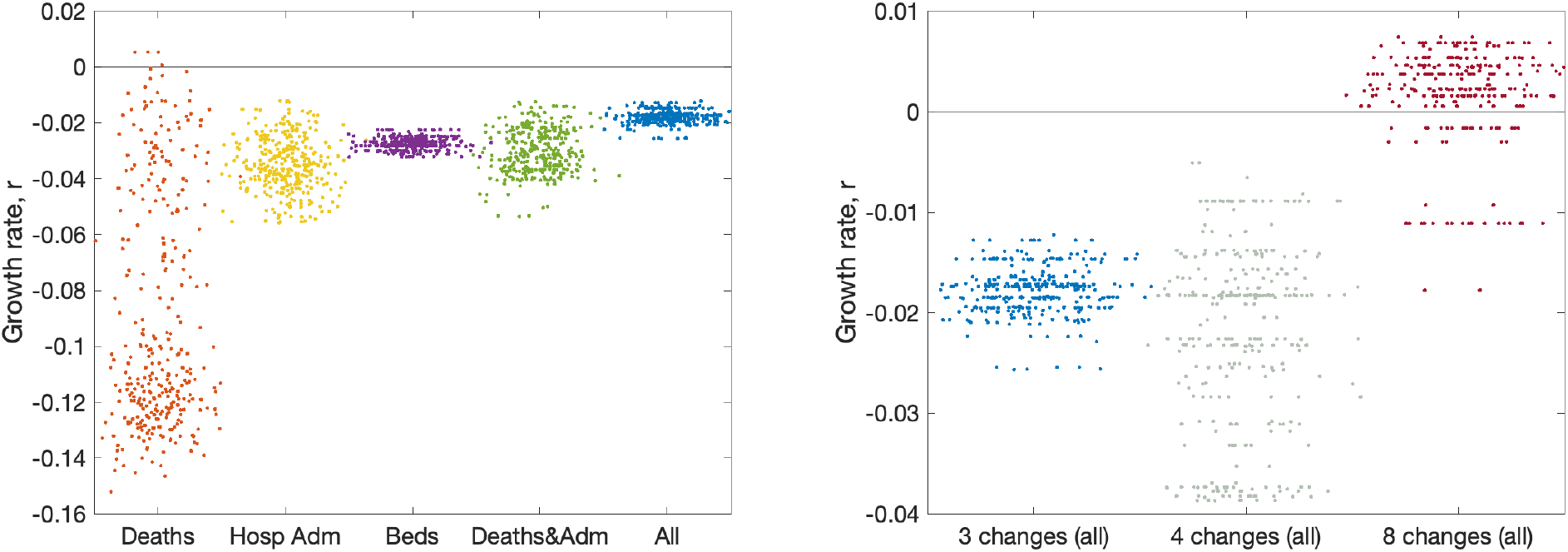
Impact of data streams and model structure on estimated growth rate. The growth rates are estimated using the predicted rate of change of new infections for London on 10th June 2020, with parameters inferred using data until 9th June 2020. The panels display posterior predictive distributions for the growth rate, where each data point corresponds to an estimate produced from a model simulation using a single parameter set sampled from the posterior distribution. To aid visualisation, we have applied a horizontal jitter to the data points. **(a)** The impact of restricting the inference to different data streams (deaths only, hospital admissions, hospital bed occupancy, deaths and admissions or all data); serology data was included in all inference. **(b)** The impact of having different numbers of lockdown phases (while using all the data); the default is three (as in Fig. 2).

In general we find that just using reported deaths produces the greatest spread of growth rates (*r*) presumably because deaths represent a small fraction of the total outbreak and therefore naturally introduce more uncertainty, and because deaths are slow to respond to dynamic changes. When hospital admissions (with or without deaths) are included in the likelihood, this generates similar predictions of the growth rate and similar levels of uncertainty in predictions. One could therefore postulate that an accurate measure of hospital admissions is the key epidemiological observable that best captures the recent growth of the epidemic.

As mentioned in Section 7, the number of phases used to describe the reduction in transmission due to lockdown has changed as the situation, model and data evolved. The model began with just two phases; before and after lockdown. However, in late May 2020, following the policy changes on 13th May 2020, we explored having three phases. Having three phases is equivalent to assuming the same level of adherence to the lockdown and social-distancing measures throughout the epidemic, with changes in transmission occurring only due to the changing policy on 23rd March 2020 and 13th May 2020. However, different number of phases can be explored (Fig. 6, right panel). Moving to four phases (with two equally spaced within the more relaxed lockdown) increases the variation, but does not have a substantial impact on the mean. Allowing eight phases (spaced every two weeks throughout lockdown) dramatically changes our estimation of the growth rate as the parameter inference responds more quickly to minor changes in observable quantities.

Lastly, it was noted in late May 2020 that one of the quantities used throughout the outbreak (number of daily hospital admissions) could lead to biased results. Hospital admissions for COVID-19 are comprised of two measures:

i. In-patients that test positive; this includes both individuals entering hospital with COVID-19 symptoms who subsequently test positive, and hospital acquired infections. Given that both of these elements feature in the hospital death data, it is difficult to separate them.
ii. Patients arriving at hospital who have previously tested positive. In the early days of the outbreak, these were individuals who had been swabbed just prior to admission; however in the latter stages there are many patients being admitted for non-COVID related problems that have previously tested positive.

It seems prudent to remove this second element from our fitting procedure, although we note that for the devolved nations this separation into in-patients and new admissions is less clear. Removing this component of admissions also means that we cannot use the number of occupied beds as part of the likelihood, as these cannot be separated by the nature of admission. In Fig. 7 we therefore compare the default fitting (used throughout this paper) with an updated method that uses in-patient admissions (together with deaths, ICU occupancy and serology when available). We observe that restricting the definition of hospital admission leads to a slight reduction in the growth rate *r* but a more pronounced reduction in the incidence.

**Fig. 7:**
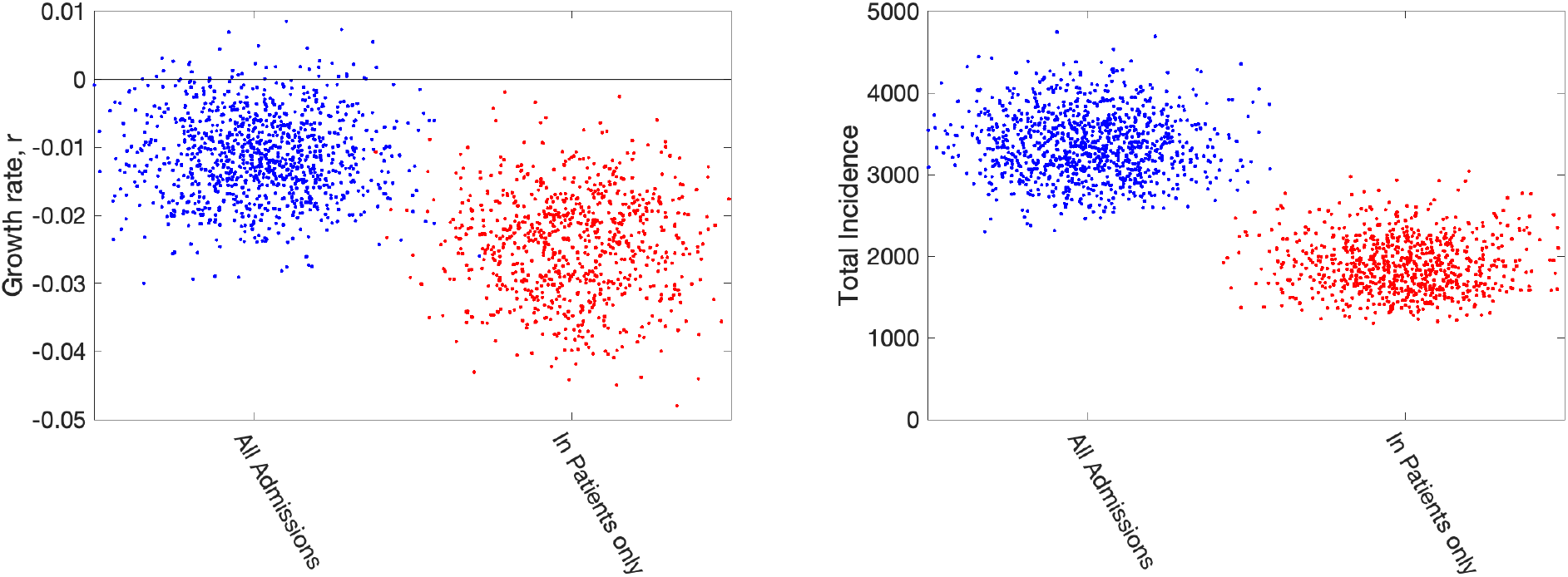
Impact of including different types of hospital admission in parameter inference. Growth rates and total incidence (asymptomatic and symptomatic) estimated from the ODE model for June 10th 2020 in London. The panels display posterior predictive distributions for the stated statistic, where each data point corresponds to an estimate produced from a model simulation using a single parameter set sampled from the posterior distribution. To aid visualisation, we have applied a horizontal jitter to the data points. In each panel, blue dots (on the left-hand side) give estimates when using all hospital admissions in the parameter inference (together with deaths, ICU occupancy and serology when available); red dots (on the right-hand side) represent estimates obtained using an alternative inference method that restricted to fitting to in-patient hospital admission data (together with deaths, ICU occupancy and serology when available). Parameters were inferred using data until 9th June 2020, while the growth rate *r* comes from the predicted rate of change of new infections.

## 9 Fits and Results at mid-June 2020

We now wish to compare how the fits made weekly (or more frequently) from late March to early June 2020 compare to later results. We note that this period also saw considerable development of the model structure as more data streams became available.

We used a fit to the data performed on 14th June 2020 (which matched to in-patient data, ICU occupancy, date of death records and serological results) to infer the change in NPIs and adherence, 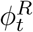, across two main intervals since lockdown: 23rd March to 13th May, and 14th May to 14th June (Fig. 8 top panels green line and shaded interval). When fed through the ODE model, the inferred distribution of parameters generate a distribution of growth rates over time (Fig. 8 bottom panels green line and shaded interval). These estimates can be compared to the estimates made at different time points for the NPI adherence and associated growth rate at that time (Fig. 8 dots and intervals). We focus on London and the North East and Yorkshire region in the main text, with other regions given in the Supplementary Material.

**Fig. 8:**
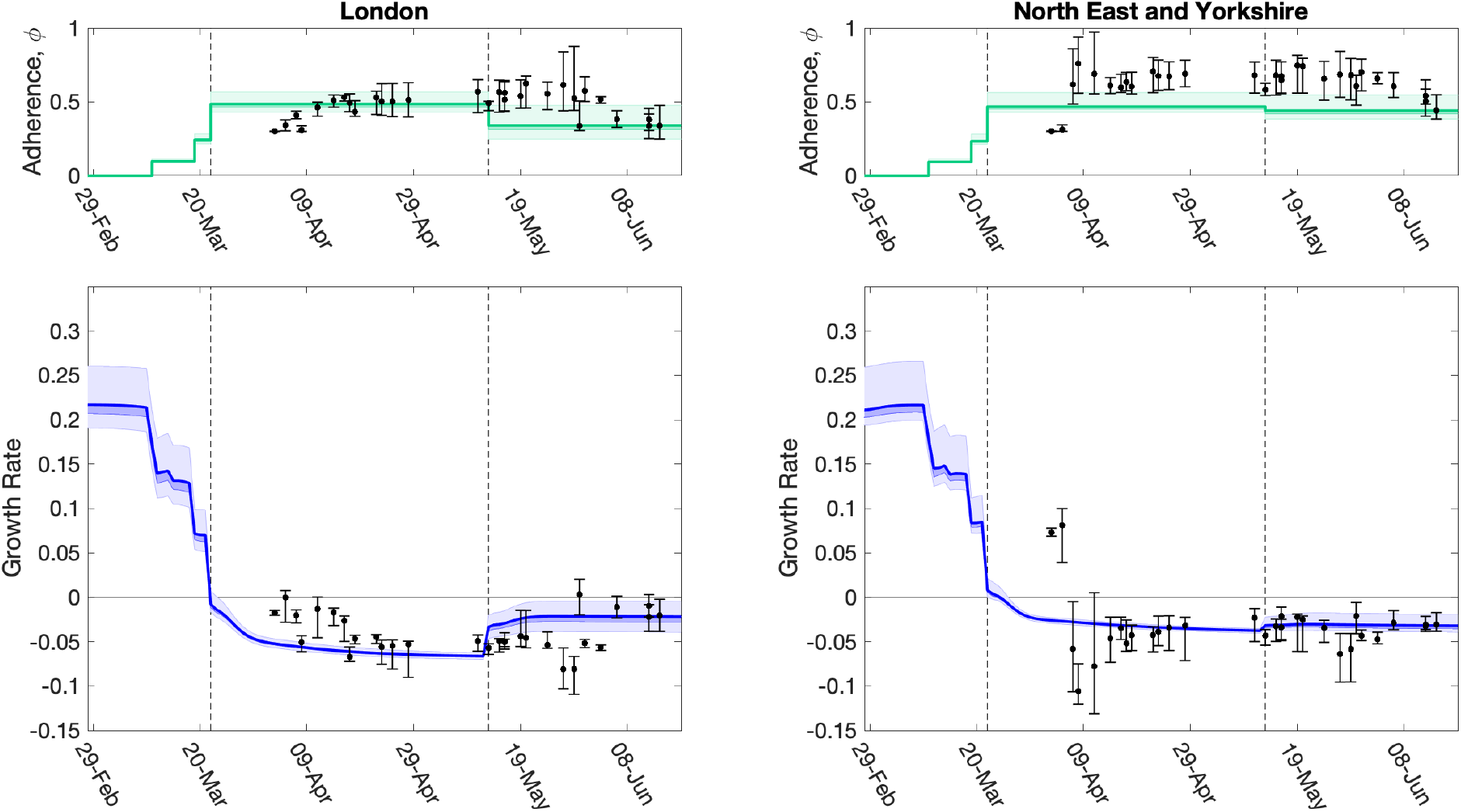
Evolution of strength of interventions and adherence values (*ϕ*_*t*_), and associated growth rate predictions (*r*) together with later model estimates. For two regions, **(left)** London, and **(right)** North East & Yorkshire, we show estimated values of the strength of interventions (comprising intervention stringency and adherence to measures, *ϕ*_*t*_), inferred from the MCMC scheme, which are translated through the model into predictions of *r*. The dots (and 95% credible intervals) show how these values have evolved over time, and are plotted for the date the MCMC inference is performed. The solid green and blue lines (together with 50% and 95% credible intervals) show our estimate of *ϕ*_*t*_ and *r* through time using a fit to the data performed on 14th June 2020 (restricting hospital data to in-patient data only). Vertical dashed lines show the two dates of main changes in policy (imposition of lockdown on 23rd March 2020, easing of restrictions on 13th May 2020), reflected in different regional *ϕ*_*t*_ values. Early changes in advice, such as social distancing, self isolation and working from home were also included in the model and their impact can be seen as early declines in the estimated growth rate *r* before 23rd March 2020.

The time profile of predicted growth rate illustrates how the imposition of lockdown measures on 23rd March 2020 led to *r* decreasing below 0. The predicted growth rate is not a step function as changes to policy precipitate changes to the age-distribution of cases which has second-order effects on *r*. The second change in *ϕ*_*t*_ (the relative strength of lockdown restrictions) on 13th May 2020 leads to an increase in *r* in all regions, although London shows one of the more pronounced increases. Despite this increase in mid-May 2020, models estimates suggest *r* remained below 0 across all regions as of 14th June 2020 (Fig. 8).

The relative strength of lockdown restrictions parameter, *ϕ*_*t*_, also captured early changes in preventative transmission behaviour that resulted from advice issued prior to the introduction of lockdown measures, such as social distancing, encouragement to work from home (from 16th March 2020) and the closure of all restaurants, pubs, cafes and schools on 20th March 2020. For all regions, we observe minor declines in the estimated growth rate following introduction of these measures, though the estimated growth rate remained above 0 (Fig. 8). As the model has evolved and the data streams become more complete, we have generally converged on the estimated growth rates from current inference. It is clear that it takes around 20 days from the time changes are enacted for them to be robustly incorporated into model parameters (see dots and 95% credible intervals in Fig. 8).

Using parameters drawn from the posterior distributions, the model produces predictive posterior distributions for multiple health outcome quantities that have a strong quantitative correspondence to the regional observations (Fig. 9). We recognise there was a looser resemblance to data on seropositivity, though salient features of the temporal profile are captured. In addition, short-term forecasts for each measure of interest have been made by continuing the simulation beyond the date of the final available data point, assuming that behaviour remains as of the final period (starting 13th May 2020).

**Fig. 9:**
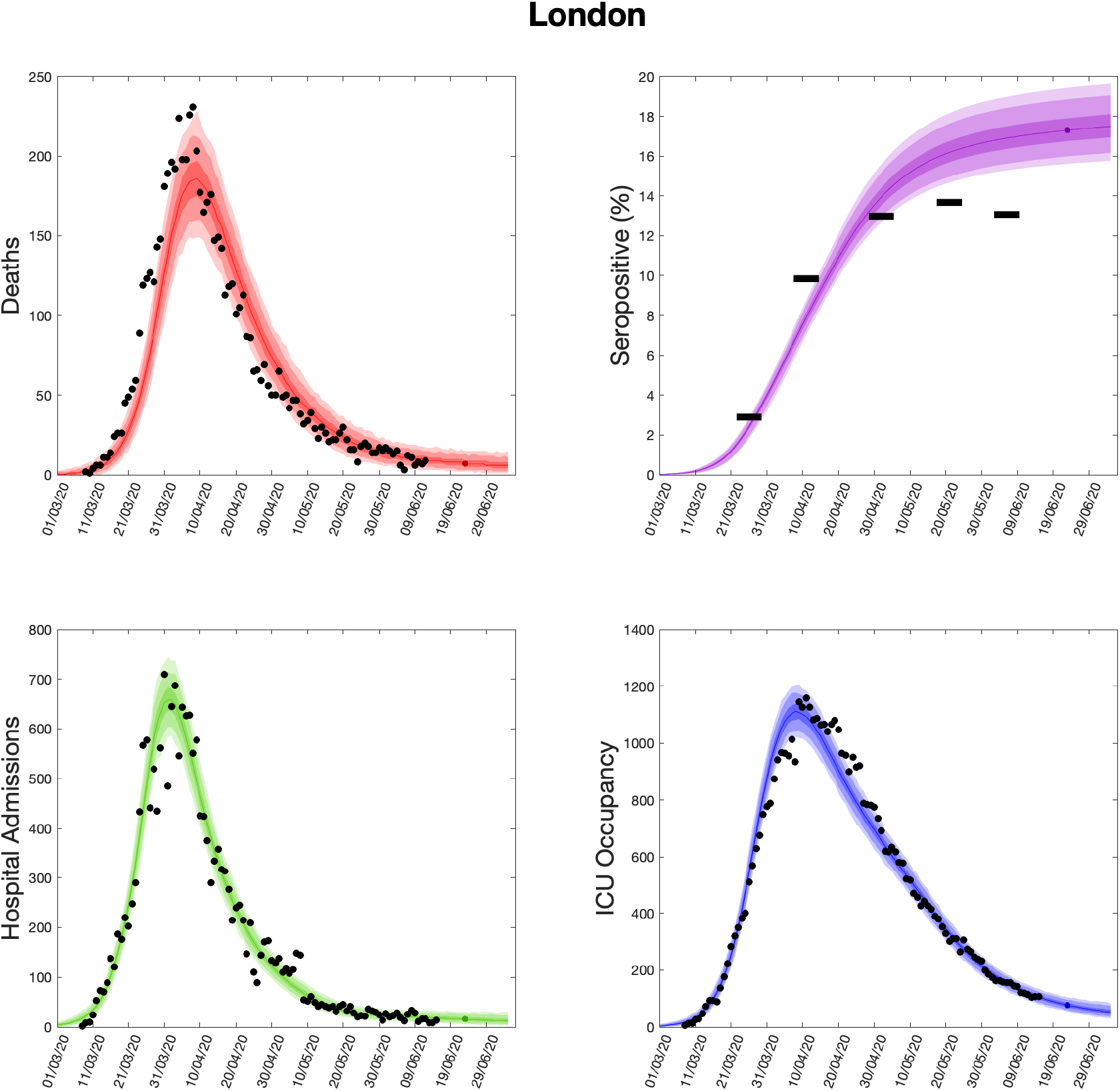
Health outcome predictions of the SARS-CoV-2 ODE transmission model from the beginning of the outbreak and three weeks into the future for London. **(Top left)** Daily deaths; **(top right)** seropositivity percentage; **(bottom left)** daily hospital admissions; **(bottom right)** ICU occupancy. In each panel: filled markers correspond to observed data, solid lines correspond to the mean outbreak over a sample of posterior parameters; shaded regions depict prediction intervals, with darker shading representing a narrower range of uncertainty (dark shading - 50%, moderate shading - 90%, light shading - 99%). The intervals represent our confidence in the fitted ODE model, and do not account for either stochastic dynamics nor the observational distribution about the deterministic predictions - which would generate far wider intervals. Predictions were produced using data up to 14th June 2020.

## 10 Discussion

In this study, we have provided an overview of the evolving MCMC inference scheme employed for calibrating the Warwick COVID-19 model [10] to the available health care, mortality and serological data streams. We have focused on the period May-June 2020, which corresponds to the first wave of the outbreak; a brief account of further refinements is given below. The work we describe was performed under extreme time pressures, working from limited initial knowledge and with data sources of varying quality. There are therefore assumptions in the model that with time and hindsight we have refined and compared to other more recently available data sources; similarly, the focus on hot-spots of infection during the summer and the rise in cases into autumn 2020 has shaped much of our methodology. This article relates the model formulation that was used to understand the dynamics, predict cases and advise policy during the first wave.

A comparison of model short-term predictions and data over time (i.e. as the outbreak has progressed) demonstrated an observable decline in the error - suggesting that our model and inference methods have improved. We have considered in some detail the choice of data sets used to infer model parameters and the impact of this choice on the key emergent properties of the growth rate *r* and the reproductive number *R*. We highlight that many of the decisions about which data sets to utilise are value judgements based on an epidemiological understanding of the relationships between disease dynamics and observed outcomes. None of the data sets available to epidemiological modellers are perfect, all have biases and delays; here we believe that by using multiple data sources in a Bayesian framework we arrive at a model that achieves a natural compromise. In particular, we have highlighted how single measures such as the number of daily deaths generate considerable uncertainty in the predicted growth rate (Fig. 6) and may be slower to identify changes in behaviour (Fig. 5). However, some questions related to the data are more fundamental; the ambiguity of what constitutes a COVID hospitalisation (Fig. 7) is shown to cause a slight difference in the estimated growth rate *r* but a more marked discrepancy in incidence.

It is important that uncertainty in the parameters governing the transmission dynamics, and its influence on predicted outcomes, be robustly conveyed. Without it, decision makers will be missing meaningful information and may assume a false sense of precision. MCMC methodologies were a suitable choice for inferring parameters in our model framework, since we were able to evaluate the likelihood function quickly enough to make the approach feasible. Nevertheless, for some model formulations and data, it may not be possible to write down or evaluate the likelihood function. In these circumstances, an alternative approach to parameter inference is via simulation-based, likelihood-free methods, such as Approximate Bayesian Computation [24–26]. Nevertheless, we recognise the appropriate mathematical structure of the model is also uncertain. Our methodology is formulated around deterministic differential equations that work well for large populations and significant levels of infection. On the other hand, stochastic effects are ignored and stochastic approaches may be needed when modelling low infection level regimes. In addition, a subset of our parameters had fixed values throughout our analyses, which means we may have underestimated the overall amount of parameter uncertainty.

As we gain collective understanding of the SARS-CoV-2 virus and the COVID-19 disease it causes, the structure of infectious disease transmission models, the inference procedure and the use of data streams to underpin these models must continuously evolve. The evolution of the model through the early phase of epidemic (up to June 2020) is documented here (Figs. 3 and 8) and we feel it is meaningful to show this evolving process rather than simply present the final finished product. A vast body of work exists describing mathematical models for different infectious outbreaks and the associated parameter inference from epidemiological data. In most cases, however, these models are fitted retrospectively, using the entire data that have been collected during an outbreak. Fitting models with such hindsight is often far more accurate than predictions made in real-time. In the case when models are deployed during active epidemics, there are also additional challenges associated with the rapid flow of detailed and accurate data. Even if robust models and methods were available from the start of an outbreak, there are still significant delays in obtaining, processing and inferring parameters from new information [5]. This is particularly crucial as new interventions are introduced or significant policy changes occur, such as the relaxation of multiple non-pharmaceutical interventions during May, June and July of 2020 or the introduction of the nationwide “test and trace” protocol [27]. Predicting the impact of such changes will inevitability be delayed by the lag between deployment and the effects on observable quantities (Fig. 5) as well as the potential need to reformulate model structure or incorporate new data streams.

Multiple refinements to the model structure and approaches have been realised since June 2020 and more are still possible. The three biggest changes have been forced by external events: the rise and spread of the Alpha (B.1.1.7) variant during the latter part of 2020; the rise and spread of the Delta (B.1.617.2) in April and May of 2021 [28]; and the development and delivery of vaccines from December 2020 onwards [29]. The two variants have necessitated an increase in the dimension of the ODEs as at least two variants need to be modelled simultaneously (either Alpha out-competing wild type, or Delta out-competing Alpha); the two new variants also require the estimation of variant specific parameters governing their relative transmission rates and the proportion of infected individuals that require hospital treatment or die from the disease [30]. The spread of these two variants is captured by looking at “S-gene failures”: the TaqPath system used to perform PCR tests in many regions of the country fails to detect the S-gene of Alpha due to a point mutation. The rise of Alpha is therefore determined by the increase of S-gene failures, while the decline of Delta is captured by the decline of S-gene failures. Vaccination also requires a large number of parameters: in particular the vaccine efficacy after one and two doses against infection, symptoms, severe illness and hospitalisation, death, and against both Alpha and Delta variants are needed within the model. We treat these additional parameters as inputs to the model, based on the estimations made by Public Health England [31].

Other changes to the model structure include using the proportion of community (known as Pillar 2) PCR samples that are positive rather than the number of positive tests. We feel that this proportion is less likely to be biased by changes in testing behaviour, and so provide a more stable estimate of the level of infection in the community. We also no-longer use serology data from blood-donors, as again this is likely to suffer from a number of confounding factors. Instead, data from the national REACT 2 study [32] is incorporated into the likelihood, and helps to anchor the total number of previously infected individuals in each region. More consideration has been given to detecting changes in the strength of social distancing (*ϕ*_*t*_). In the original model *ϕ*_*t*_ was inferred in two main phases: the main lockdown (from 23rd March to 13th May 2020) and the more relaxed restrictions (from 13th May 2020). In practice, there will be continuous changes to this quantity as the population’s behaviour varies (not necessarily in response to government guidelines), given the importance to public health planning of rapidly detecting such changes we now estimate the values of *ϕ*_*t*_ on a weekly timescale but assume the value to only vary slowly (unless there has been a major change to the restrictions). Finally, we have assumed that many of the observable epidemiological quantities (such as hospitalisation and death) are related in a fixed way to the age-distribution of infection in the population. In reality, the medical treatment of COVID-19 cases in the UK has changed dramatically since the first few cases in early March 2020, such that the risk of mortality, the need for hospitalisation and the duration of hospital stay have all changed. Such changes have been incorporated periodically into the model structure, informed from hospital data sources.

Despite all of these improvements over the last year, there are still aspects that could be further improved. The understanding that infection may be partially driven by nosocomial transmission [33, 34], while significant mortality is due to infection in care homes [35, 36], suggests that additional compartments capturing these components could greatly improve model realism if the necessary data were available throughout the course of the epidemic. Similarly, schools, universities and some workplaces pose additional risks, so there is merit in considering how these amplifiers of community infection could be incorporated within the general framework [37–39]. Additionally, if in a regime with much lower levels of infection in the community, it may be prudent to adopt a stochastic model formulation at a finer spatial resolution to capture localised outbreak clusters, although the potential heterogeneity in local parameters may preclude accurate prediction at this scale.

In summary, if epidemiological models are to be used as part of the scientific discussion of controlling a disease outbreak it is vital that these models capture current biological understanding and are continually matched to all available data in real time. Our work on COVID-19 presented here highlights some of the challenges with predicting a novel outbreak in a rapidly changing environment. Probably the greatest weakness is the time that it inevitably takes to respond – both in terms of developing the appropriate model and inference structure, and the mechanisms to process any data sources, but also in terms of delay between real-world changes and their detection within any inference scheme. Both of these can be shortened by well-informed preparations; having the necessary suite of models supported by the latest most efficient inference techniques could be hugely beneficial when rapid and robust predictive results are required.

## Data Availability

This work uses data provided by patients and collected by the NHS as part of their care and support #DataSavesLives. We are extremely grateful to the 2,648 frontline NHS clinical and research staff and volunteer medical students, who collected this data in challenging circumstances; and the generosity of the participants and their families for their individual contributions in these difficult times. The CO-CIN data was collated by ISARIC4C Investigators. ISARIC4C welcomes applications for data and material access through our Independent Data and Material Access Committee (https://isaric4c.net).
Data on cases were obtained from the COVID-19 Hospitalisation in England Surveillance System (CHESS) data set that collects detailed data on patients infected with COVID-19. Data on COVID-19 deaths were obtained from Public Health England. These data contain confidential information, with public data deposition non-permissible for socioeconomic reasons. The CHESS data resides with the National Health Service (www.nhs.gov.uk) whilst the death data are available from Public Health England (www.phe.gov.uk).

## Author contributions

**Conceptualisation:** Matt J. Keeling.

**Data curation:** Matt J. Keeling; Glen Guyver-Fletcher; Alexander Holmes.

**CO-CIN Data provision:** Malcolm G. Semple and the ISARIC4C Investigators.

**Formal analysis:** Matt J. Keeling.

**Investigation:** Matt J. Keeling.

**Methodology:** Matt J. Keeling.

**Software:** Matt J. Keeling; Edward M. Hill; Louise Dyson; Michael J. Tildesley.

**Validation:** Matt J. Keeling; Edward M. Hill; Louise Dyson; Michael J. Tildesley.

**Visualisation:** Matt J. Keeling.

**Writing - original draft:** Matt J. Keeling; Michael J. Tildesley; Edward M. Hill; Louise Dyson.

**Writing - review & editing:** Matt J. Keeling; Edward M. Hill; Glen Guyver-Fletcher; Alexander Holmes; Malcolm G. Semple; Louise Dyson; Michael J. Tildesley.

## Patient and public involvement

This was an urgent public health research study in response to a Public Health Emergency of International Concern. Patients or the public were not involved in the design, conduct, or reporting of this rapid response research.

## Ethical considerations

Ethical approval was given by the South Central - Oxford C Research Ethics Committee in England (Ref 13/SC/0149), the Scotland A Research Ethics Committee (Ref 20/SS/0028), and the WHO Ethics Review Committee (RPC571 and RPC572, 25 April 2013).

Data from the CHESS database were supplied after anonymisation under strict data protection protocols agreed between the University of Warwick and Public Health England. The ethics of the use of these data for these purposes was agreed by Public Health England with the Government’s SPI-M(O) / SAGE committees.

## Data availability

This work uses data provided by patients and collected by the NHS as part of their care and support #DataSavesLives. We are extremely grateful to the 2,648 frontline NHS clinical and research staff and volunteer medical students, who collected this data in challenging circumstances; and the generosity of the participants and their families for their individual contributions in these difficult times. The CO-CIN data was collated by ISARIC4C Investigators. ISARIC4C welcomes applications for data and material access through our Independent Data and Material Access Committee (https://isaric4c.net).

Data on cases were obtained from the COVID-19 Hospitalisation in England Surveillance System (CHESS) data set that collects detailed data on patients infected with COVID-19. Data on COVID-19 deaths were obtained from Public Health England. These data contain confidential information, with public data deposition non-permissible for socioeconomic reasons. The CHESS data resides with the National Health Service (www.nhs.gov.uk) whilst the death data are available from Public Health England (www.phe.gov.uk).

## Acknowledgements

We acknowledge the support of Jeremy J Farrar, Nahoko Shindo, Devika Dixit, Nipunie Rajapakse, Piero Olliaro, Lyndsey Castle, Martha Buckley, Debbie Malden, Katherine Newell, Kwame O’Neill, Emmanuelle Denis, Claire Petersen, Scott Mullaney, Sue MacFarlane, Chris Jones, Nicole Maziere, Katie Bullock, Emily Cass, William Reynolds, Milton Ashworth, Ben Catterall, Louise Cooper, Terry Foster, Paul Matthew Ridley, Anthony Evans, Catherine Hartley, Chris Dunn, Debby Sales, Diane Latawiec, Erwan Trochu, Eve Wilcock, Innocent Gerald Asiimwe, Isabel Garcia-Dorival, J. Eunice Zhang, Jack Pilgrim, Jane A Armstrong, Jordan J. Clark, Jordan Thomas, Katharine King, Katie Alexandra Ahmed, Krishanthi S Subramaniam, Lauren Lett, Laurence McEvoy, Libby van Tonder, Lucia Alicia Livoti, Nahida S Miah, Rebecca K. Shears, Rebecca Louise Jensen, Rebekah Penrice-Randal, Robyn Kiy, Samantha Leanne Barlow, Shadia Khandaker, Soeren Metelmann, Tessa Prince, Trevor R Jones, Benjamin Brennan, Agnieska Szemiel, Siddharth Bakshi, Daniella Lefteri, Maria Mancini, Julien Martinez, Angela Elliott, Joyce Mitchell, John McLauchlan, Aislynn Taggart, Oslem Dincarslan, Annette Lake, Claire Petersen, and Scott Mullaney.

## ISARIC 4C Investigators

Consortium Lead Investigator: J Kenneth Baillie.

Chief Investigator: Malcolm G Semple.

Co-Lead Investigator: Peter JM Openshaw.

ISARIC Clinical Coordinator: Gail Carson.

Co-Investigators: Beatrice Alex, Benjamin Bach, Wendy S Barclay, Debby Bogaert, Meera Chand, Graham S Cooke, Annemarie B Docherty, Jake Dunning, Ana da Silva Filipe, Tom Fletcher, Christopher A Green, Ewen M Harrison, Julian A Hiscox, Antonia Ying Wai Ho, Peter W Horby, Samreen Ijaz, Saye Khoo, Paul Klenerman, Andrew Law, Wei Shen Lim, Alexander, J Mentzer, Laura Merson, Alison M Meynert, Mahdad Noursadeghi, Shona C Moore, Massimo Palmarini, William A Paxton, Georgios Pollakis, Nicholas Price, Andrew Rambaut, David L Robertson, Clark D Russell, Vanessa Sancho-Shimizu, Janet T Scott, Louise Sigfrid, Tom Solomon, Shiranee Sriskandan, David Stuart, Charlotte Summers, Richard S Tedder, Emma C Thomson, Ryan S Thwaites, Lance CW Turtle, Maria Zambon. Project Managers Hayley Hardwick, Chloe Donohue, Jane Ewins, Wilna Oosthuyzen, Fiona Griffiths. Data Analysts: Lisa Norman, Riinu Pius, Tom M Drake, Cameron J Fairfield, Stephen Knight, Kenneth A Mclean, Derek Murphy, Catherine A Shaw. Data and Information System Manager: Jo Dalton, Michelle Girvan, Egle Saviciute, Stephanie Roberts Janet Harrison, Laura Marsh, Marie Connor. Data integration and presentation: Gary Leeming, Andrew Law, Ross Hendry. Material Management: William Greenhalf, Victoria Shaw, Sarah McDonald. Outbreak Laboratory Volunteers: Katie A. Ahmed, Jane A Armstrong, Milton Ashworth, Innocent G Asiimwe, Siddharth Bakshi, Samantha L Barlow, Laura Booth, Benjamin Brennan, Katie Bullock, Benjamin WA Catterall, Jordan J Clark, Emily A Clarke, Sarah Cole, Louise Cooper, Helen Cox, Christopher Davis, Oslem Dincarslan, Chris Dunn, Philip Dyer, Angela Elliott, Anthony Evans, Lewis WS Fisher, Terry Foster, Isabel Garcia-Dorival, Willliam Greenhalf, Philip Gunning, Catherine Hartley, Antonia Ho, Rebecca L Jensen, Christopher B Jones, Trevor R Jones, Shadia Khandaker, Katharine King, Robyn T. Kiy, Chrysa Koukorava, Annette Lake, Suzannah Lant, Diane Latawiec, L Lavelle-Langham, Daniella Lefteri, Lauren Lett, Lucia A Livoti, Maria Mancini, Sarah McDonald, Laurence McEvoy, John McLauchlan, Soeren Metelmann, Nahida S Miah, Joanna Middleton, Joyce Mitchell, Shona C Moore, Ellen G Murphy, Rebekah Penrice-Randal, Jack Pilgrim, Tessa Prince, Will Reynolds, P. Matthew Ridley, Debby Sales, Victoria E Shaw, Rebecca K Shears, Benjamin Small, Krishanthi S Subramaniam, Agnieska Szemiel, Aislynn Taggart, Jolanta Tanianis, Jordan Thomas, Erwan Trochu, Libby van Tonder, Eve Wilcock, J. Eunice Zhang. Local Principal Investigators: Kayode Adeniji, Daniel Agranoff, Ken Agwuh, Dhiraj Ail, Ana Alegria, Brian Angus, Abdul Ashish, Dougal Atkinson, Shahedal Bari, Gavin Barlow, Stella Barnass, Nicholas Barrett, Christopher Bassford, David Baxter, Michael Beadsworth, Jolanta Bernatoniene, John Berridge, Nicola Best, Pieter Bothma, David Brealey, Robin Brittain-Long, Naomi Bulteel, Tom Burden, Andrew Burtenshaw, Vikki Caruth, David Chadwick, Duncan Chambler, Nigel Chee, Jenny Child, Srikanth Chukkambotla, Tom Clark, Paul Collini, Catherine Cosgrove, Jason Cupitt, Maria-Teresa Cutino-Moguel, Paul Dark, Chris Dawson, Samir Dervisevic, Phil Donnison, Sam Douthwaite, Ingrid DuRand, Ahilanadan Dushianthan, Tristan Dyer, Cariad Evans, Chi Eziefula, Chrisopher Fegan, Adam Finn, Duncan Fullerton, Sanjeev Garg, Sanjeev Garg, Atul Garg, Jo Godden, Arthur Goldsmith, Clive Graham, Elaine Hardy, Stuart Hartshorn, Daniel Harvey, Peter Havalda, Daniel B Hawcutt, Maria Hobrok, Luke Hodgson, Anita Holme, Anil Hormis, Michael Jacobs, Susan Jain, Paul Jennings, Agilan Kaliappan, Vidya Kasipandian, Stephen Kegg, Michael Kelsey, Jason Kendall, Caroline Kerrison, Ian Kerslake, Oliver Koch, Gouri Koduri, George Koshy, Shondipon Laha, Susan Larkin, Tamas Leiner, Patrick Lillie, James Limb, Vanessa Linnett, Jeff Little, Michael MacMahon, Emily MacNaughton, Ravish Mankregod, Huw Masson, Elijah Matovu, Katherine McCullough, Ruth McEwen, Manjula Meda, Gary Mills, Jane Minton, Mariyam Mirfenderesky, Kavya Mohandas, Quen Mok, James Moon, Elinoor Moore, Patrick Morgan, Craig Morris, Katherine Mortimore, Samuel Moses, Mbiye Mpenge, Rohinton Mulla, Michael Murphy, Megan Nagel, Thapas Nagarajan, Mark Nelson, Igor Otahal, Mark Pais, Selva Panchatsharam, Hassan Paraiso, Brij Patel, Justin Pepperell, Mark Peters, Mandeep Phull, Stefania Pintus, Jagtur Singh Pooni, Frank Post, David Price, Rachel Prout, Nikolas Rae, Henrik Reschreiter, Tim Reynolds, Neil Richardson, Mark Roberts, Devender Roberts, Alistair Rose, Guy Rousseau, Brendan Ryan, Taranprit Saluja, Aarti Shah, Prad Shanmuga, Anil Sharma, Anna Shawcross, Jeremy Sizer, Richard Smith, Catherine Snelson, Nick Spittle, Nikki Staines, Tom Stambach, Richard Stewart, Pradeep Subudhi, Tamas Szakmany, Kate Tatham, Jo Thomas, Chris Thompson, Robert Thompson, Ascanio Tridente, Darell Tupper - Carey, Mary Twagira, Andrew Ustianowski, Nick Vallotton, Lisa Vincent-Smith, Shico Visuvanathan, Alan Vuylsteke, Sam Waddy, Rachel Wake, Andrew Walden, Ingeborg Welters, Tony Whitehouse, Paul Whittaker, Ashley Whittington, Meme Wijesinghe, Martin Williams, Lawrence Wilson, Sarah Wilson, Stephen Winchester, Martin Wiselka, Adam Wolverson, Daniel G Wooton, Andrew Workman, Bryan Yates, Peter Young.

## SUPPLEMENTARY MATERIAL

**Fig. S1:**
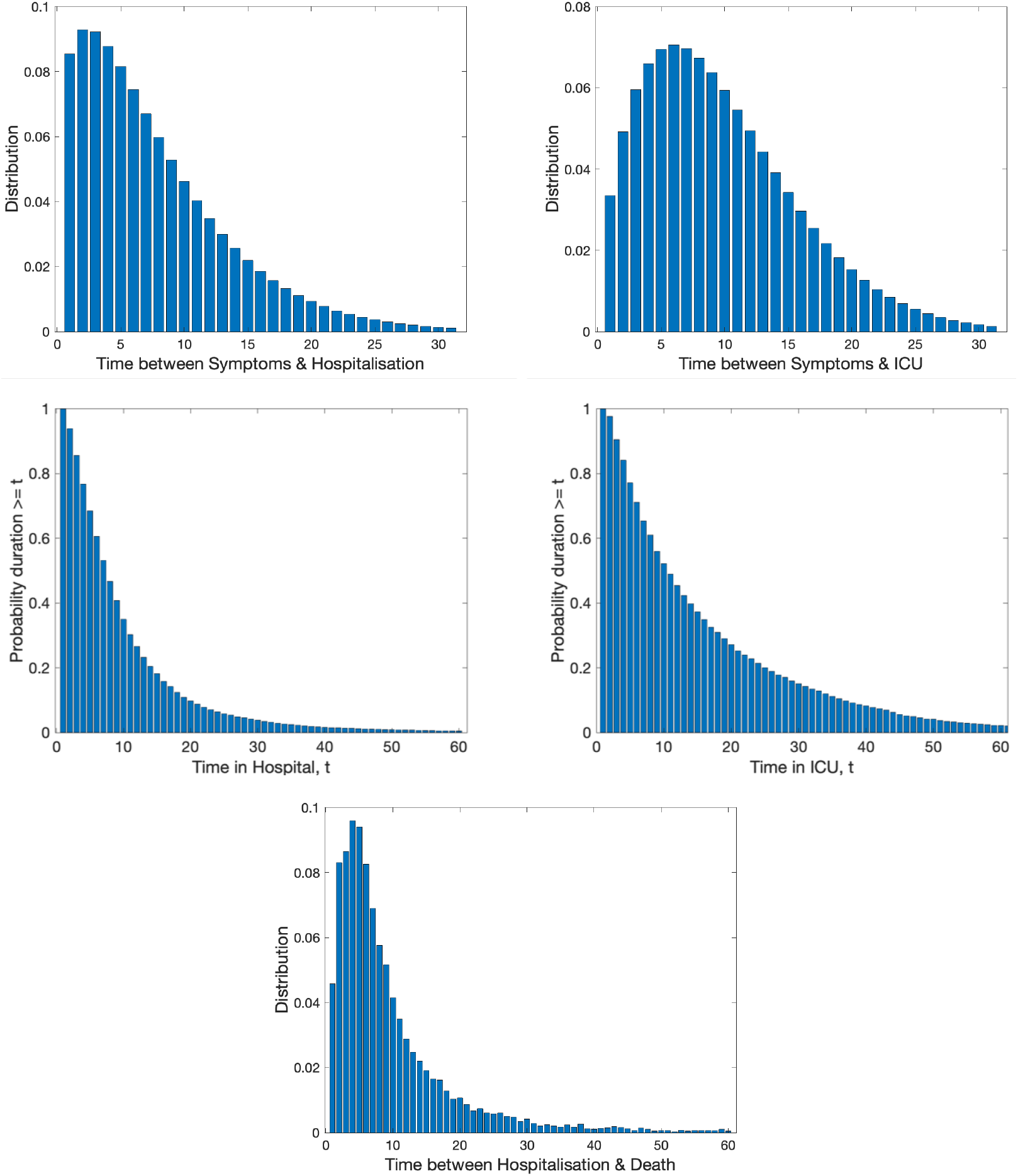
Distribution of times and delays used in the model. (Top row, left) Time between symptom onset and hospitalisation, conditional on hospital admission occurring. (Top row, right) Time between symptom onset and ICU admittance, conditional on ICU admission occurring. We modelled these two delays as Weibull distributions estimated from the available data: Time between symptom onset and hospitalisation, *λ* = 8.4, *k* = 1.4; Time between symptom onset and ICU admittance, *λ* = 10.9, *k* = 1.7. We did not take these distributions directly from the available data which was relatively sparse due to the lack of certainty involving the date of symptom onset for many patients. (Middle row, left) Probability of the length of stay in hospital equalling or exceeding a duration of *t* days. (Middle row, right) Probability of the length of stay in ICU equalling or exceeding a duration of *t* days. (Bottom row) Time between hospitalisation and death. The remaining three distributions, displayed in the middle and bottoms rows, we took directly from the available data. All distributions are based on individual patient data as recorded by the COVID-19 Hospitalisation in England Surveillance System (CHESS) [21] and the ISARIC WHO Clinical Characterisation Protocol UK (CCP-UK) database sourced from the COVID-19 Clinical Information Network (CO-CIN) [22, 23].

**Fig. S2:**
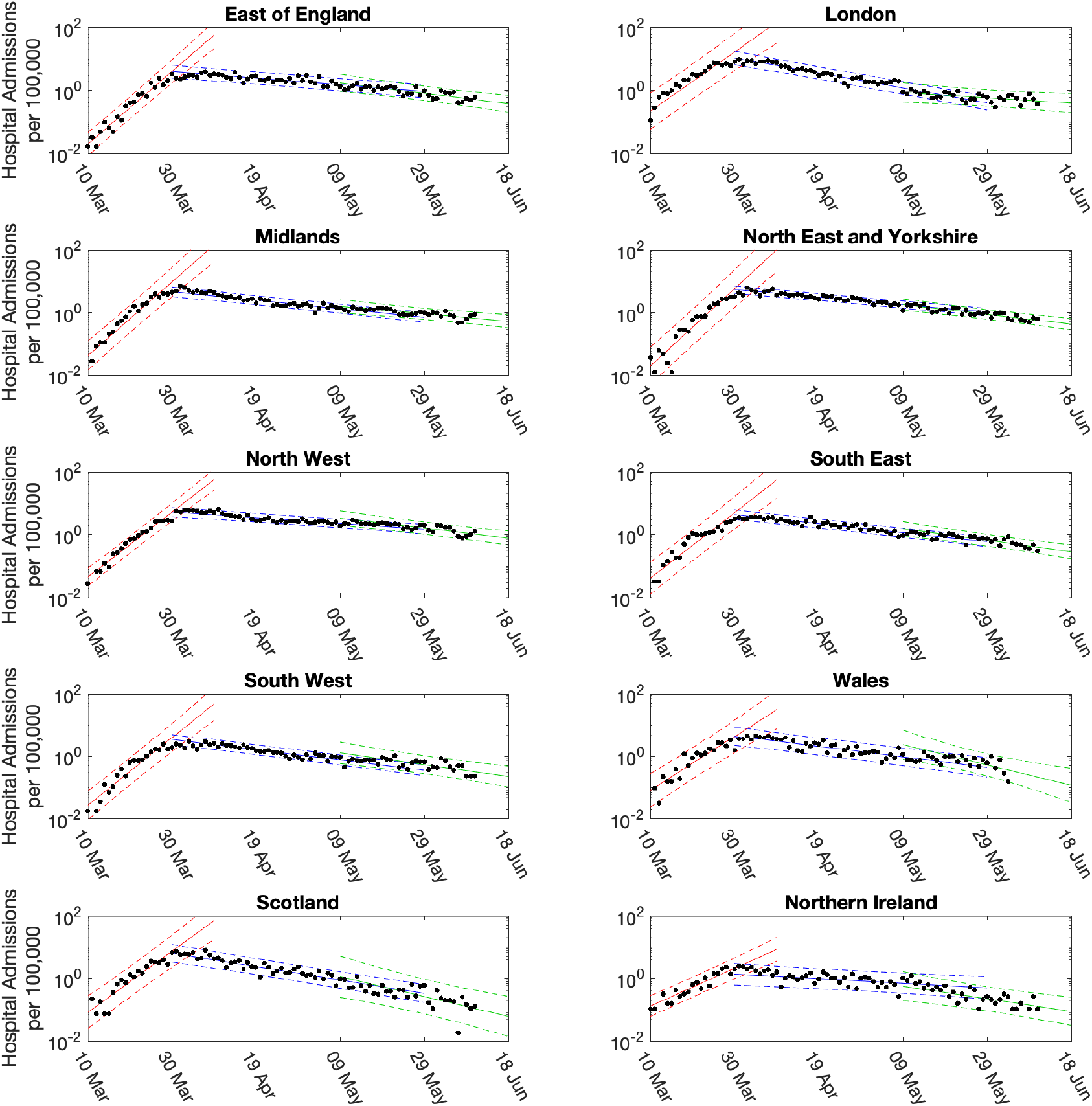
Linear fits to daily hospital admissions per 100,000 individuals in each region. Points show the number of daily admissions to hospital (both in-patients testing positive and patients entering hospital following a positive test); results are plotted on a log scale. Three simple fits to the data are shown for pre-lockdown (red), strict-lockdown (blue) and relaxed-lockdown phases (green). Fits are a limit linear fit to the logged data (mean estimates depicted by solid lines, 95% confidence intervals by the dashed lines).

**Fig. S3:**
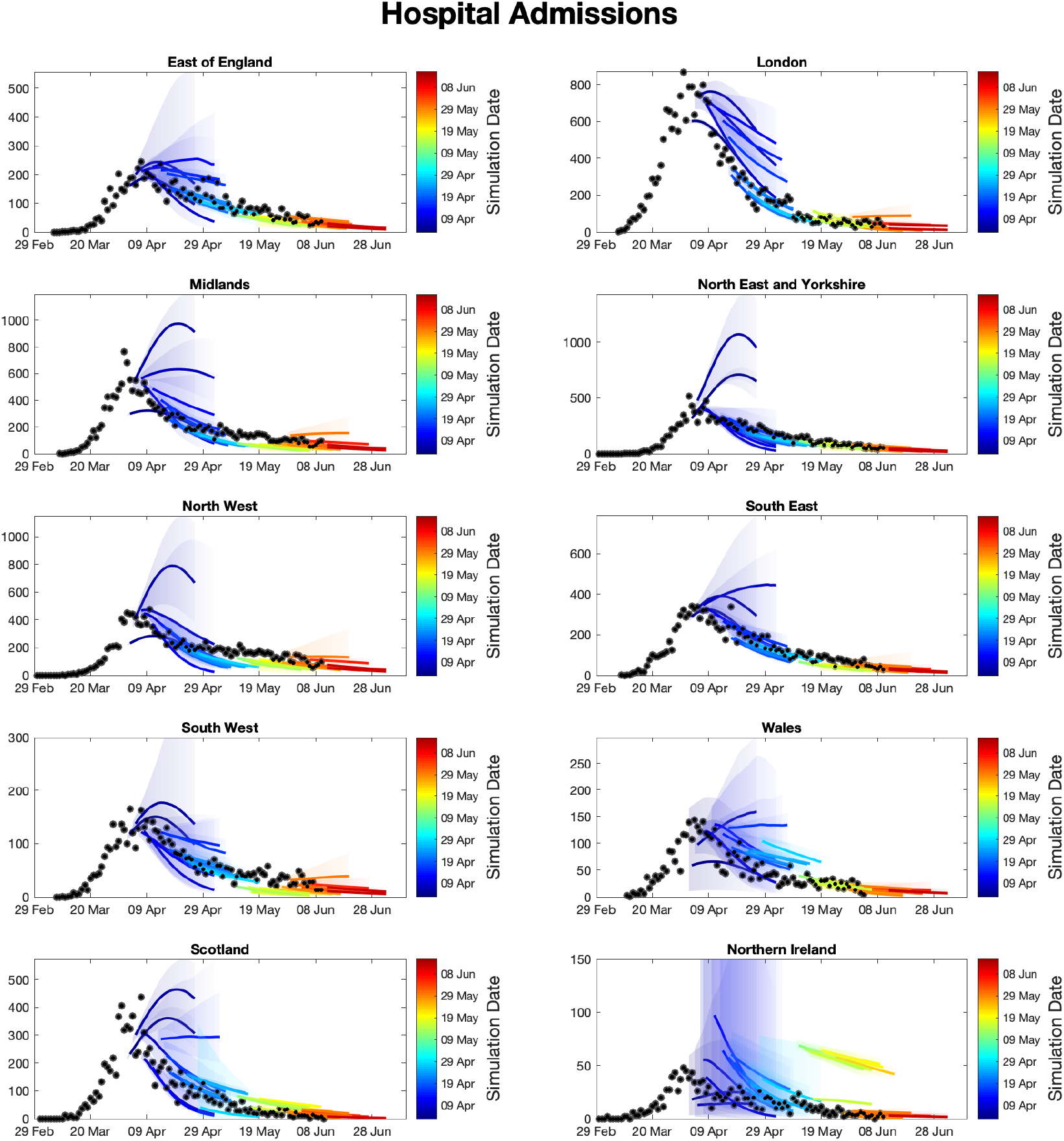
Sequential comparison of daily hospital admission model results and data in each region. For all daily hospital admissions with COVID-19 in each region, we show the raw data (black dots) and a set of short-term predictions generated at different points during the outbreak. Changes to model fit reflect both improvements in model structure as well as increased amounts of data.

**Fig. S4:**
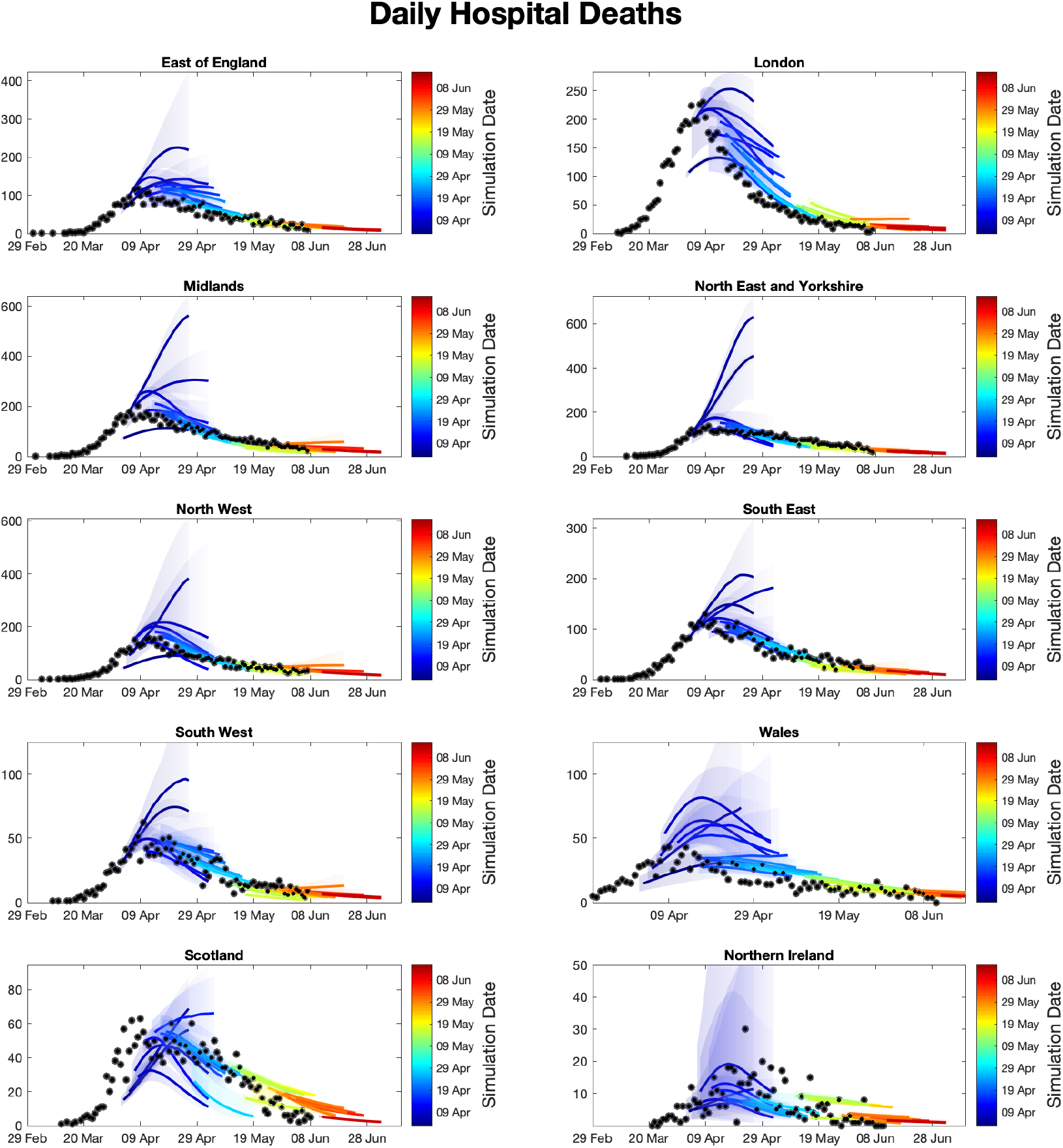
Sequential comparison of daily hospital death model results and data in each region. For all daily hospital deaths with COVID-19 in each region, we show the raw data (black dots) and a set of short-term predictions generated at different points during the outbreak. Changes to model fit reflect both improvements in model structure as well as increased amounts of data.

**Fig. S5:**
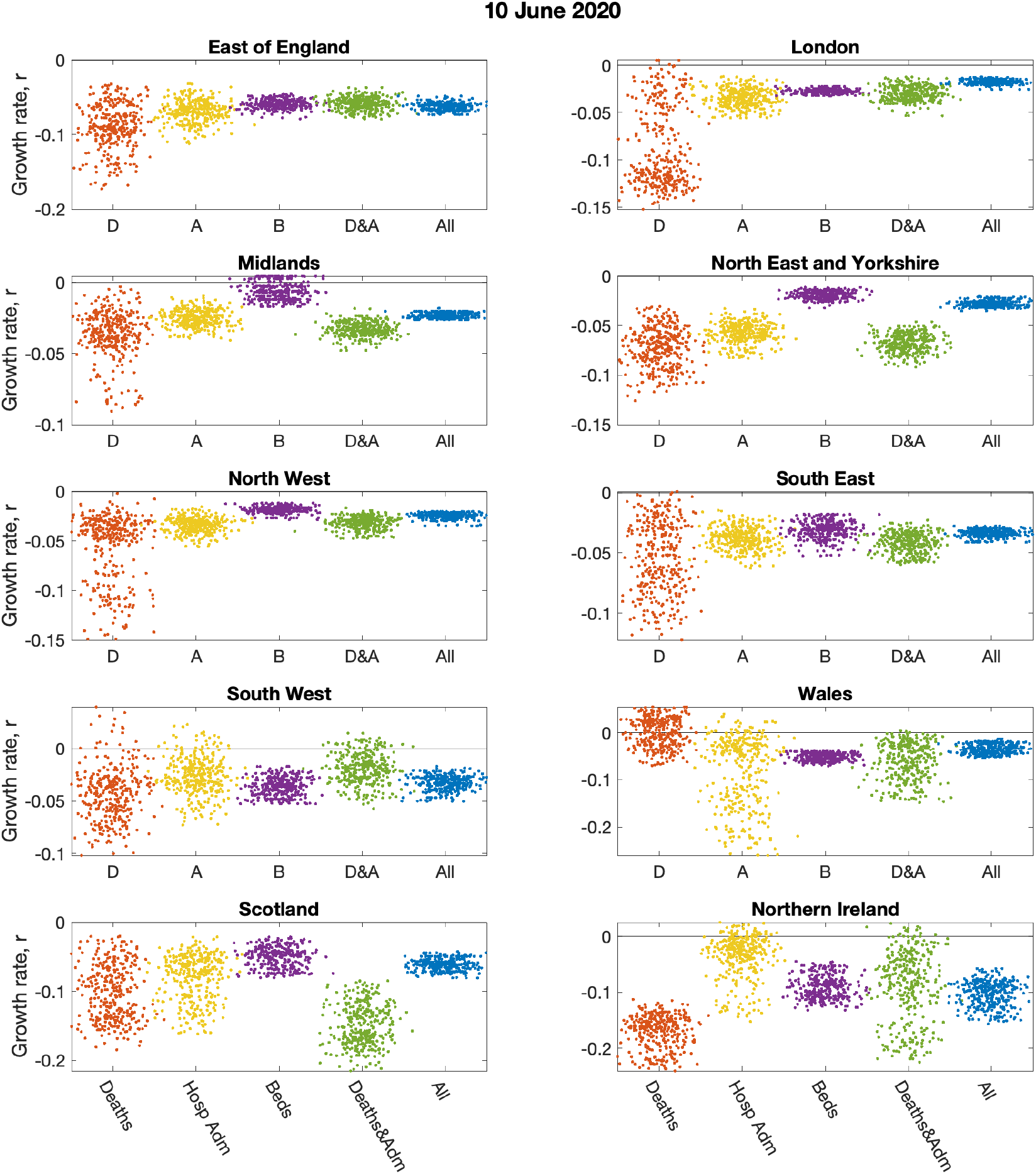
Impact of data streams on estimated growth rates, by region. The impact on the regional growth rate (estimated from the ODE epidemic on 10th June 2020) of restricting the inference to different data streams (deaths only, hospital admissions, hospital bed occupancy, deaths and admissions or all data); the serology data was included in all inference. Parameters were inferred using data until 9th June 2020, while the *r* value comes from the change in predicted rate of change of new cases.

**Fig. S6:**
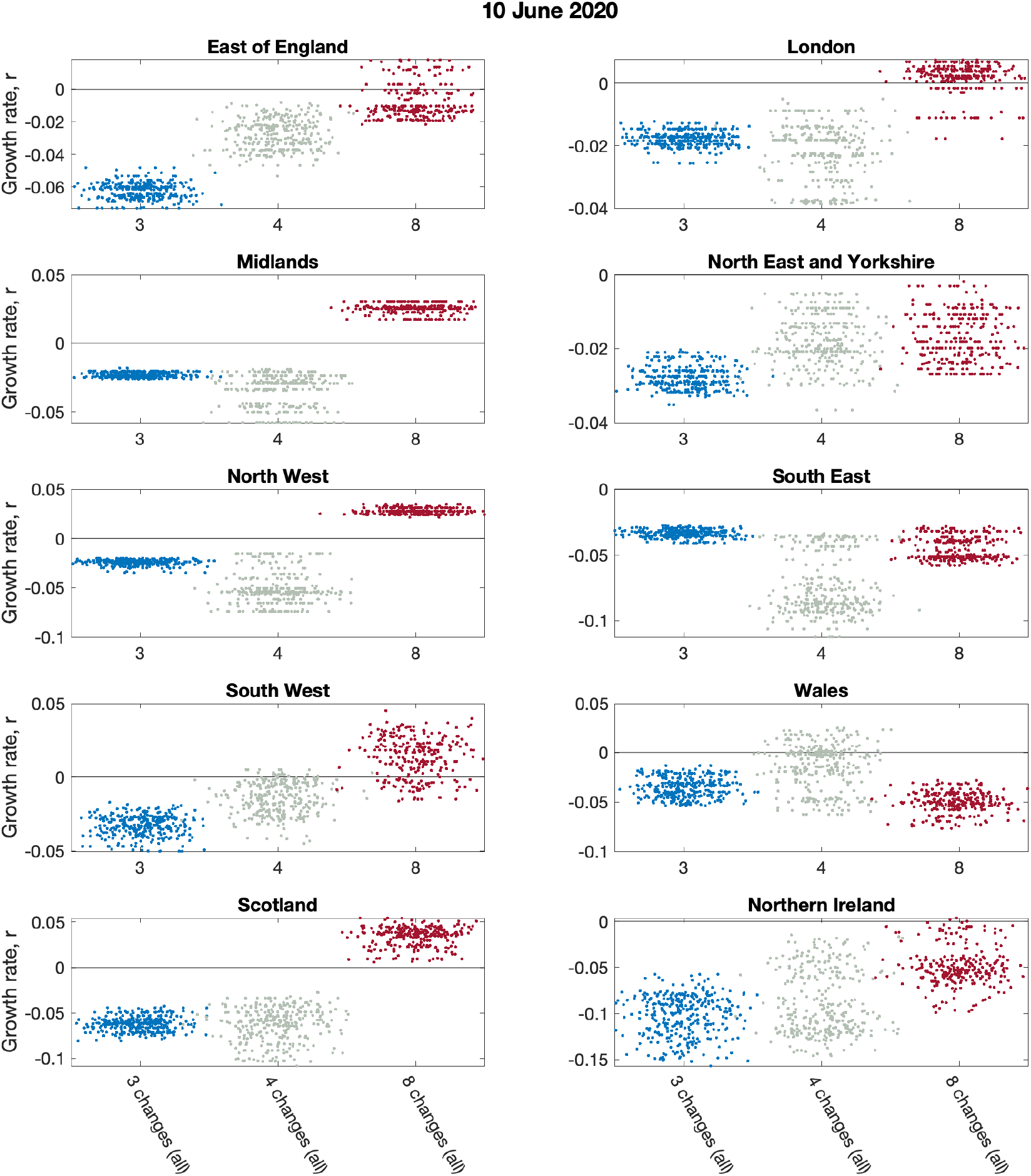
Impact of model structure on estimated growth rates, by region. Analysis of having different numbers of lockdown phases on the estimated regional growth rate on 10th June 2020, while using all the data. The number of lockdown phases tested were there (blue dots), four (green dots) and eight (red dots). Parameters were inferred using data until 9th June 2020, while the *r* value comes from the change in predicted rate of change of new cases.

**Fig. S7:**
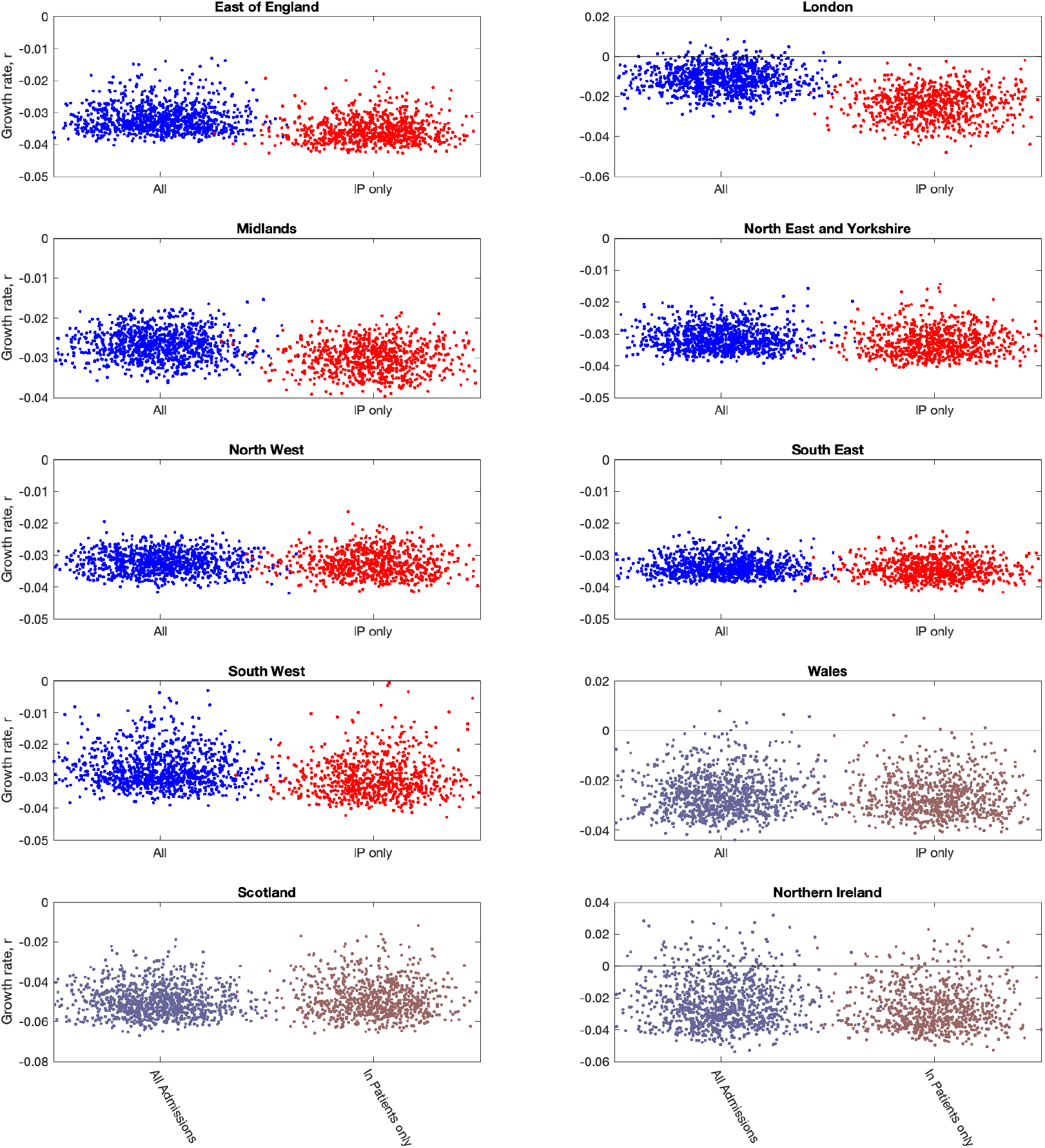
Impact of including different types of hospital admission in parameter inference on the estimated regional growth rates (on 10th June 2020). For each region, growth rates were estimated from the ODE epidemic for 10th June 2020. In each panel, blue dots (on the left-hand side) give *r* estimates when using all hospital admissions in the parameter inference (together with deaths, ICU occupancy and serology when available); red dots (on the right-hand side) represent *r* estimates using an alternative inference method that restricted to fitting to in-patient hospital admission data (together with deaths, ICU occupancy and serology when available). Parameters were inferred using data until 9th June 2020, while the *r* value comes from the change in predicted rate of change of new cases. We observe that restricting the definition of hospital admission leads to a slight reduction in the growth rate *r* but a more pronounced reduction in the incidence. (This separation is not possible for the devolved nations, but the associated distributions are shown for completeness.)

**Fig. S8:**
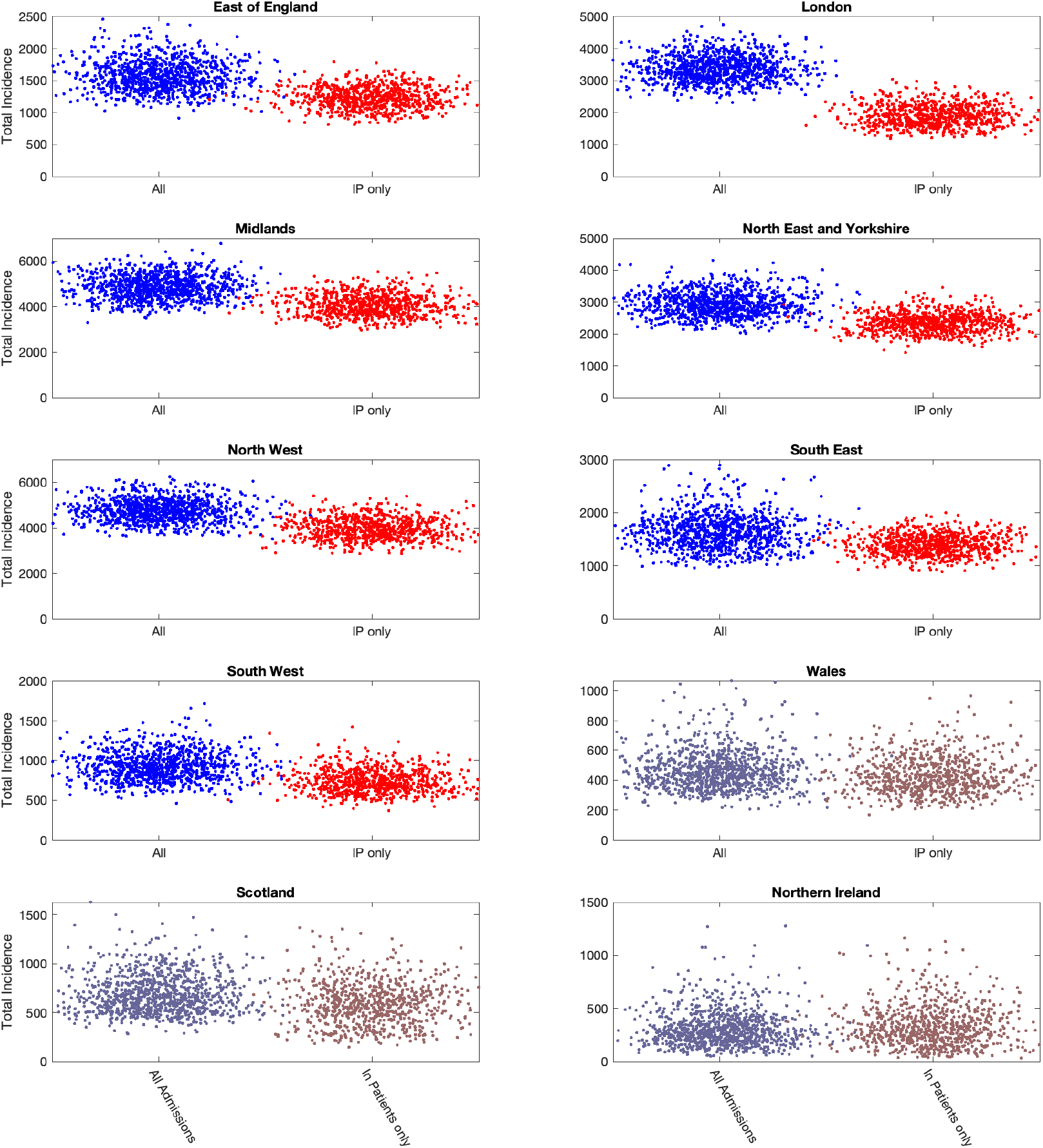
Impact of including different types of hospital admission in parameter inference on the daily incidence (on 10th June 2020). For each region, daily incidences were estimated from the ODE epidemic for 10th June 2020. In each panel, blue dots (on the left-hand side) give incidence estimates when using all hospital admissions in the parameter inference (together with deaths, ICU occupancy and serology when available); red dots (on the right-hand side) represent incidence estimates using an alternative inference method that restricted to fitting to in-patient hospital admission data (together with deaths, ICU occupancy and serology when available). Parameters were inferred using data until 9th June 2020. We observe that restricting the definition of hospital admission leads to a pronounced reduction in the incidence. (This separation is not possible for the devolved nations, but the associated distributions are shown for completeness.)

**Fig. S9:**
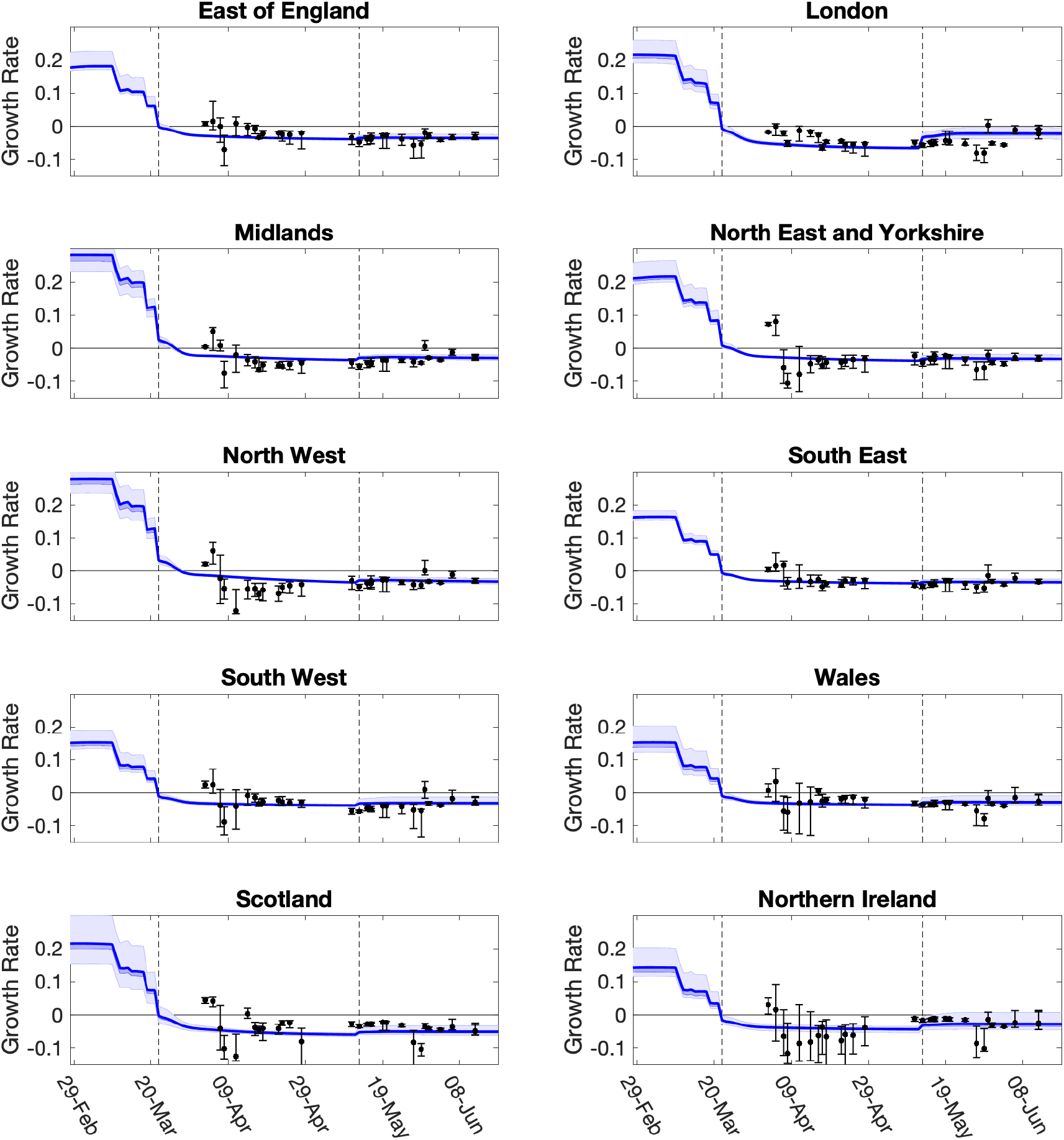
Evolution of growth rate predictions and most recent model estimates in each of the ten regions. For each region, we show how predictions of *r* have evolved over time (dots and 95% credible intervals). These predictions are from the date the MCMC inference is performed. The solid blue line (together with 50% and 95% credible intervals) shows our estimate of *r* through time using the most recent fit to the data (performed on 14th June 2020 using in-patient data only). Vertical dashed lines show the two dates of main changes in policy, reflected in different regional *ϕ* values.

**Fig. S10:**
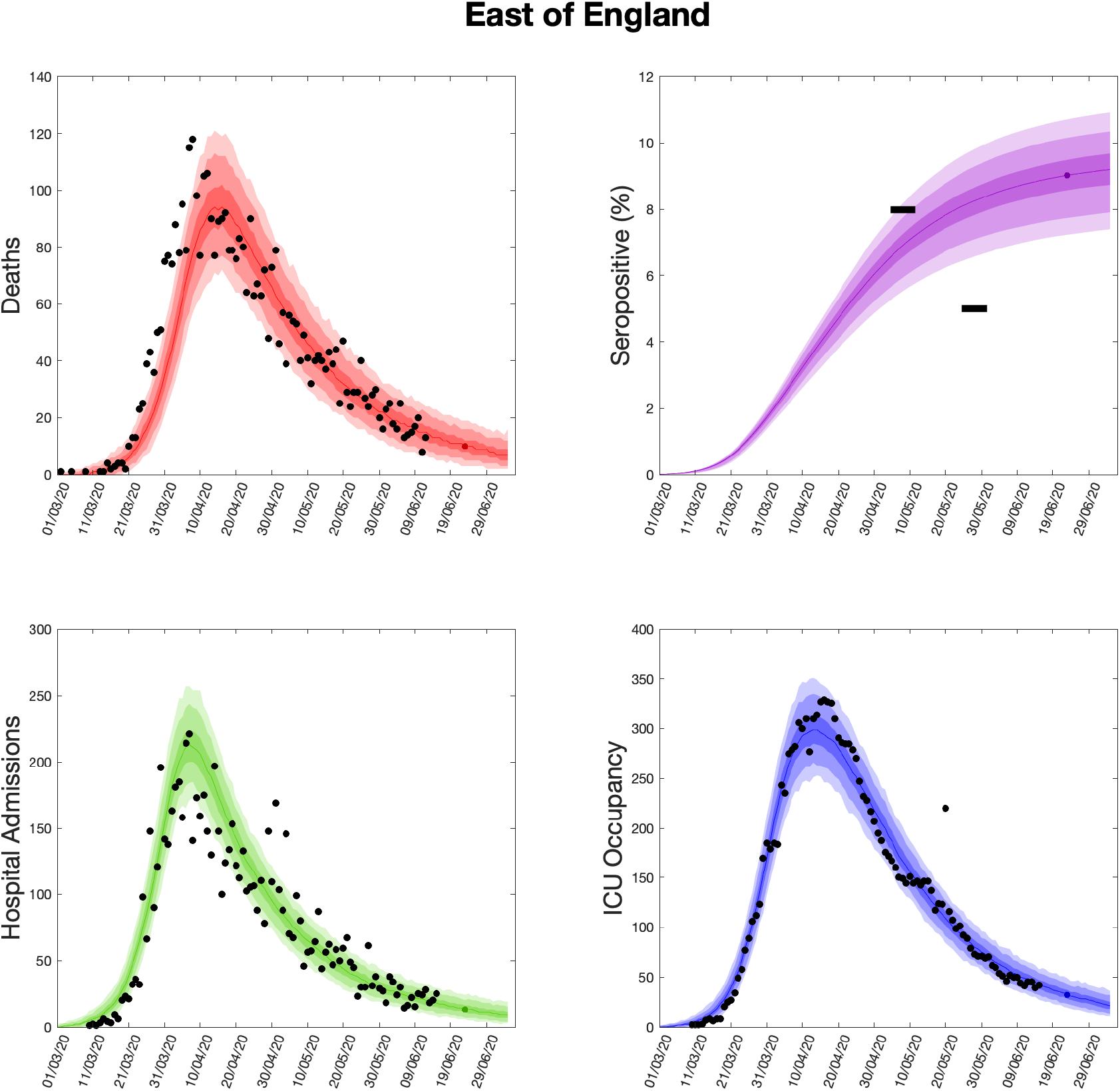
Health outcome predictions of the ODE from the beginning of the outbreak and 3 weeks into the future for the East of England region. **(Top left)** Daily deaths; **(top right)** seropositivity percentage; **(bottom left)** daily hospital admissions; **(bottom right)** ICU occupancy. In each panel: filled markers correspond to observed data, solid lines correspond to the mean outbreak over a sample of posterior parameters; shaded regions depict prediction intervals, with darker shading representing a narrower range of uncertainty (dark shading - 50%, moderate shading - 90%, light shading - 99%). The intervals represent our confidence in the fitted ODE model, and do not account for either stochastic dynamics nor the observational distribution about the deterministic predictions - which would generate far wider intervals. Predictions were produced using data up to 14th June 2020.

**Fig. S11:**
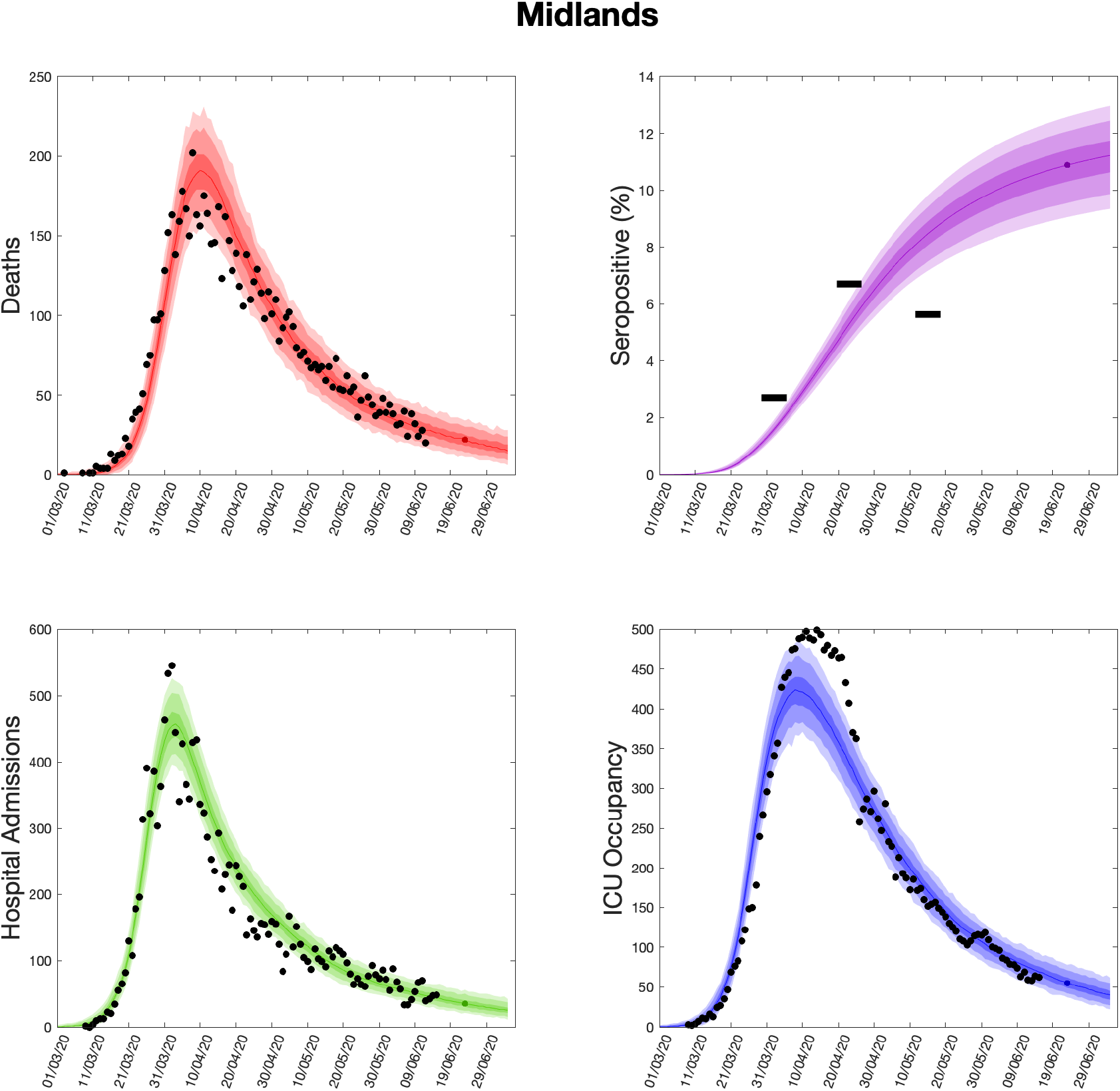
Health outcome predictions of the ODE from the beginning of the outbreak and 3 weeks into the future for the Midlands region. **(Top left)** Daily deaths; **(top right)** seropositivity percentage; **(bottom left)** daily hospital admissions; **(bottom right)** ICU occupancy. In each panel: filled markers correspond to observed data, solid lines correspond to the mean outbreak over a sample of posterior parameters; shaded regions depict prediction intervals, with darker shading representing a narrower range of uncertainty (dark shading - 50%, moderate shading - 90%, light shading - 99%). The intervals represent our confidence in the fitted ODE model, and do not account for either stochastic dynamics nor the observational distribution about the deterministic predictions - which would generate far wider intervals. Predictions were produced using data up to 14th June 2020.

**Fig. S12:**
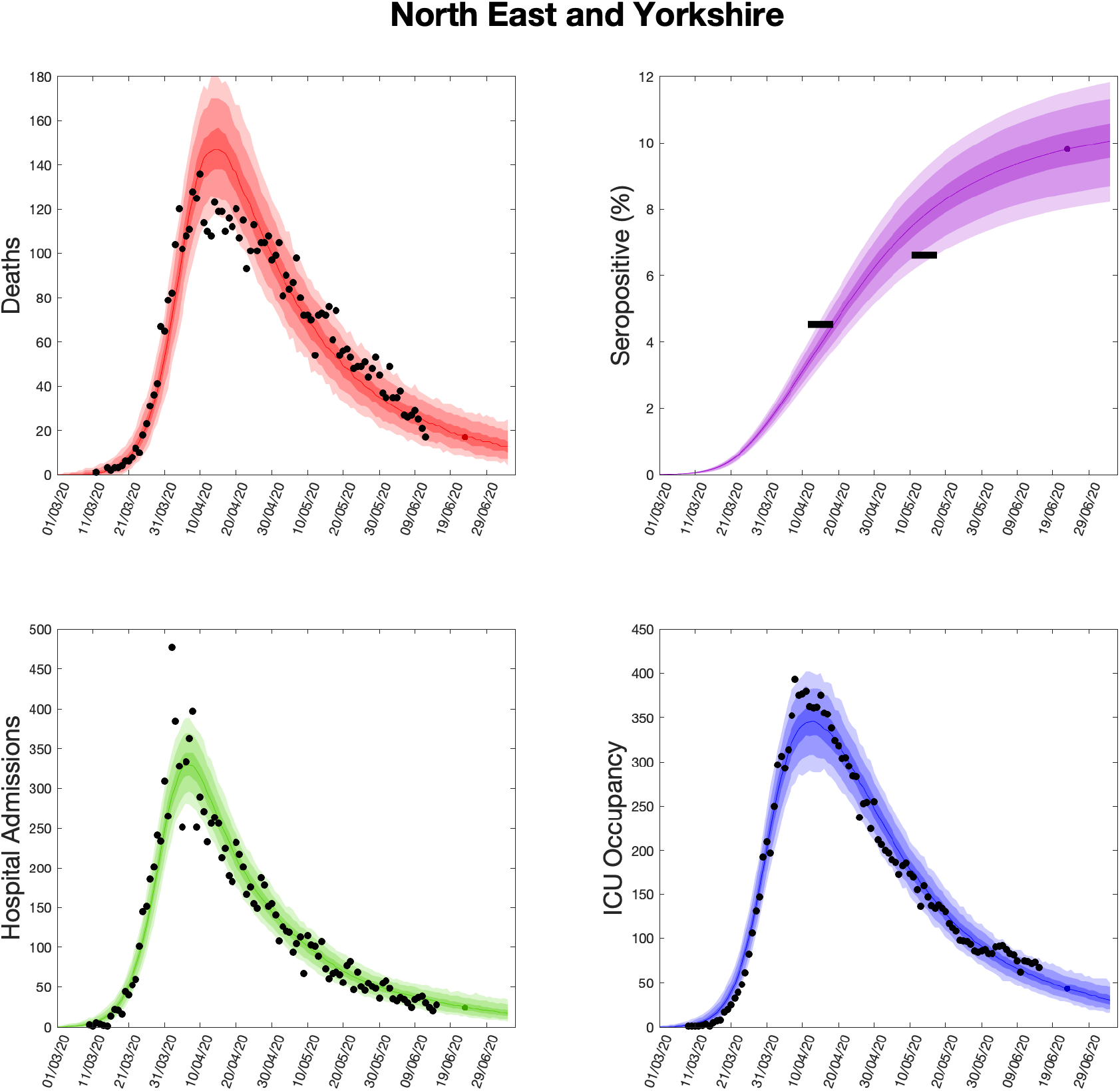
Health outcome predictions of the ODE from the beginning of the outbreak and 3 weeks into the future for the North East & Yorkshire region. **(Top left)** Daily deaths; **(top right)** seropositivity percentage; **(bottom left)** daily hospital admissions; **(bottom right)** ICU occupancy. In each panel: filled markers correspond to observed data, solid lines correspond to the mean outbreak over a sample of posterior parameters; shaded regions depict prediction intervals, with darker shading representing a narrower range of uncertainty (dark shading - 50%, moderate shading - 90%, light shading - 99%). The intervals represent our confidence in the fitted ODE model, and do not account for either stochastic dynamics nor the observational distribution about the deterministic predictions - which would generate far wider intervals. Predictions were produced using data up to 14th June 2020.

**Fig. S13:**
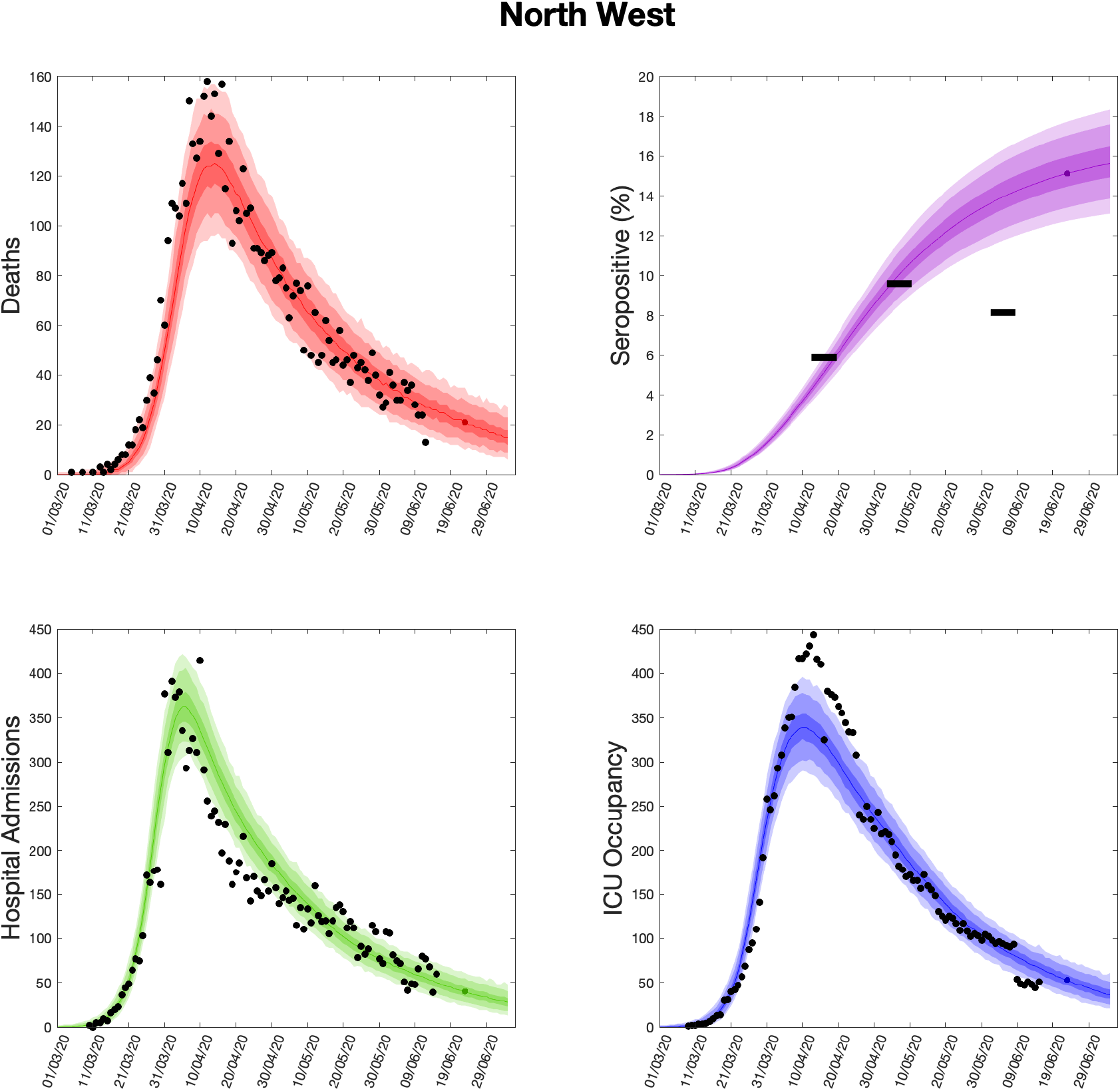
Health outcome predictions of the ODE from the beginning of the outbreak and 3 weeks into the future for the North West region. **(Top left)** Daily deaths; **(top right)** seropositivity percentage; **(bottom left)** daily hospital admissions; **(bottom right)** ICU occupancy. In each panel: filled markers correspond to observed data, solid lines correspond to the mean outbreak over a sample of posterior parameters; shaded regions depict prediction intervals, with darker shading representing a narrower range of uncertainty (dark shading - 50%, moderate shading - 90%, light shading - 99%). The intervals represent our confidence in the fitted ODE model, and do not account for either stochastic dynamics nor the observational distribution about the deterministic predictions - which would generate far wider intervals. Predictions were produced using data up to 14th June 2020.

**Fig. S14:**
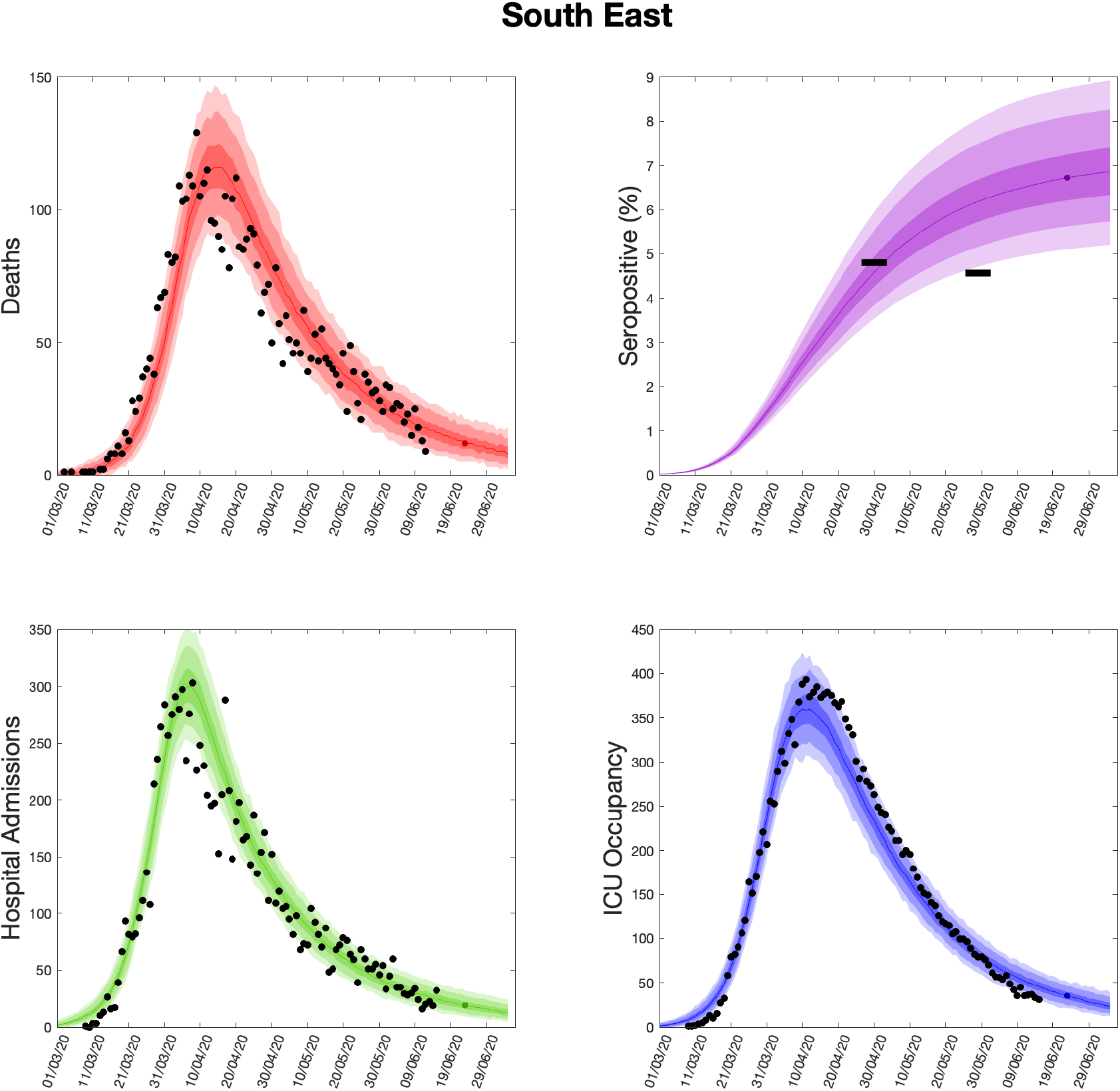
Health outcome predictions of the ODE from the beginning of the outbreak and 3 weeks into the future for the South East region. **(Top left)** Daily deaths; **(top right)** seropositivity percentage; **(bottom left)** daily hospital admissions; **(bottom right)** ICU occupancy. In each panel: filled markers correspond to observed data, solid lines correspond to the mean outbreak over a sample of posterior parameters; shaded regions depict prediction intervals, with darker shading representing a narrower range of uncertainty (dark shading - 50%, moderate shading - 90%, light shading - 99%). The intervals represent our confidence in the fitted ODE model, and do not account for either stochastic dynamics nor the observational distribution about the deterministic predictions - which would generate far wider intervals. Predictions were produced using data up to 14th June 2020.

**Fig. S15:**
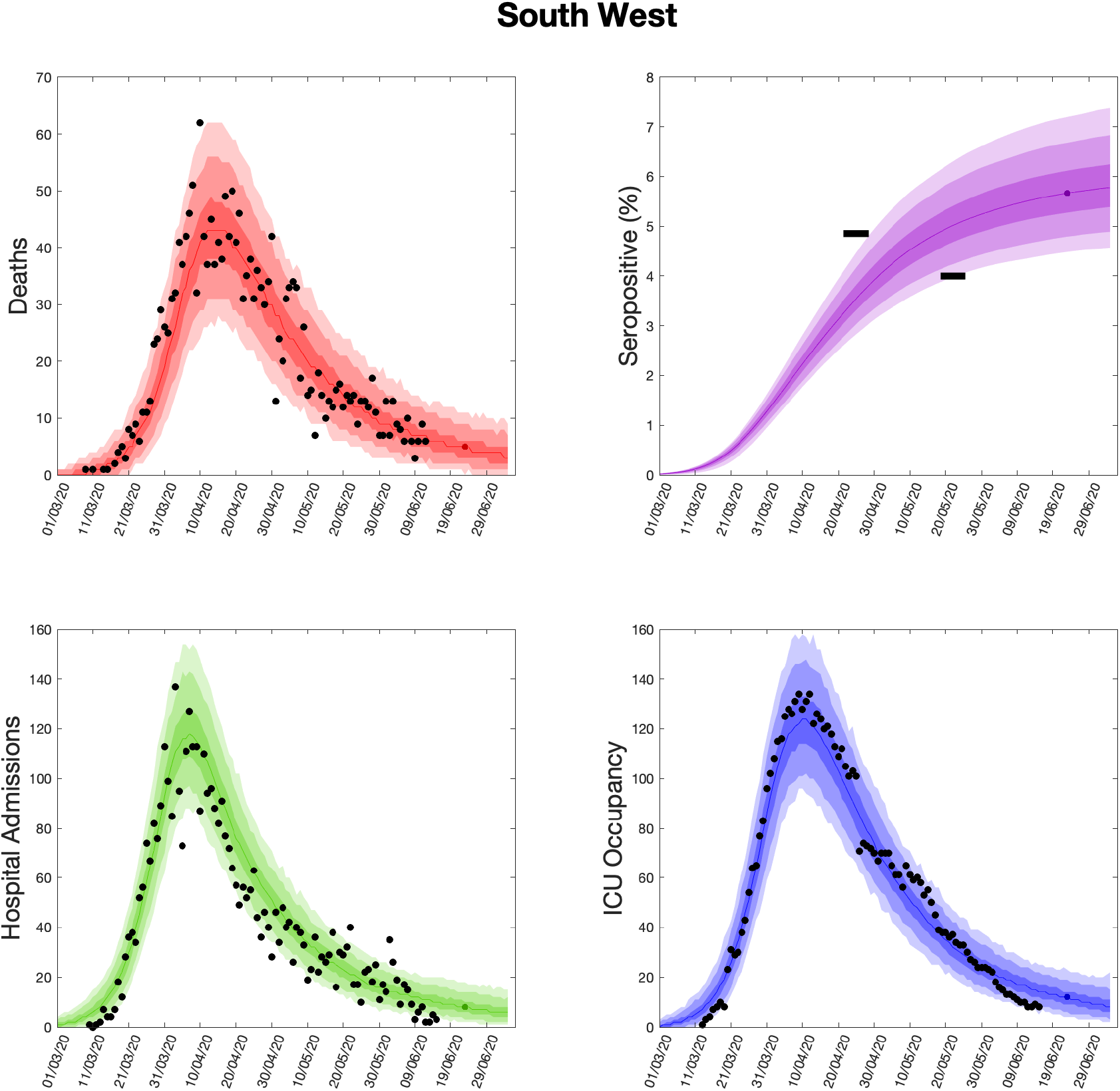
Health outcome predictions of the ODE from the beginning of the outbreak and 3 weeks into the future for the South West region. **(Top left)** Daily deaths; **(top right)** seropositivity percentage; **(bottom left)** daily hospital admissions; **(bottom right)** ICU occupancy. In each panel: filled markers correspond to observed data, solid lines correspond to the mean outbreak over a sample of posterior parameters; shaded regions depict prediction intervals, with darker shading representing a narrower range of uncertainty (dark shading - 50%, moderate shading - 90%, light shading - 99%). The intervals represent our confidence in the fitted ODE model, and do not account for either stochastic dynamics nor the observational distribution about the deterministic predictions - which would generate far wider intervals. Predictions were produced using data up to 14th June 2020.

**Fig. S16:**
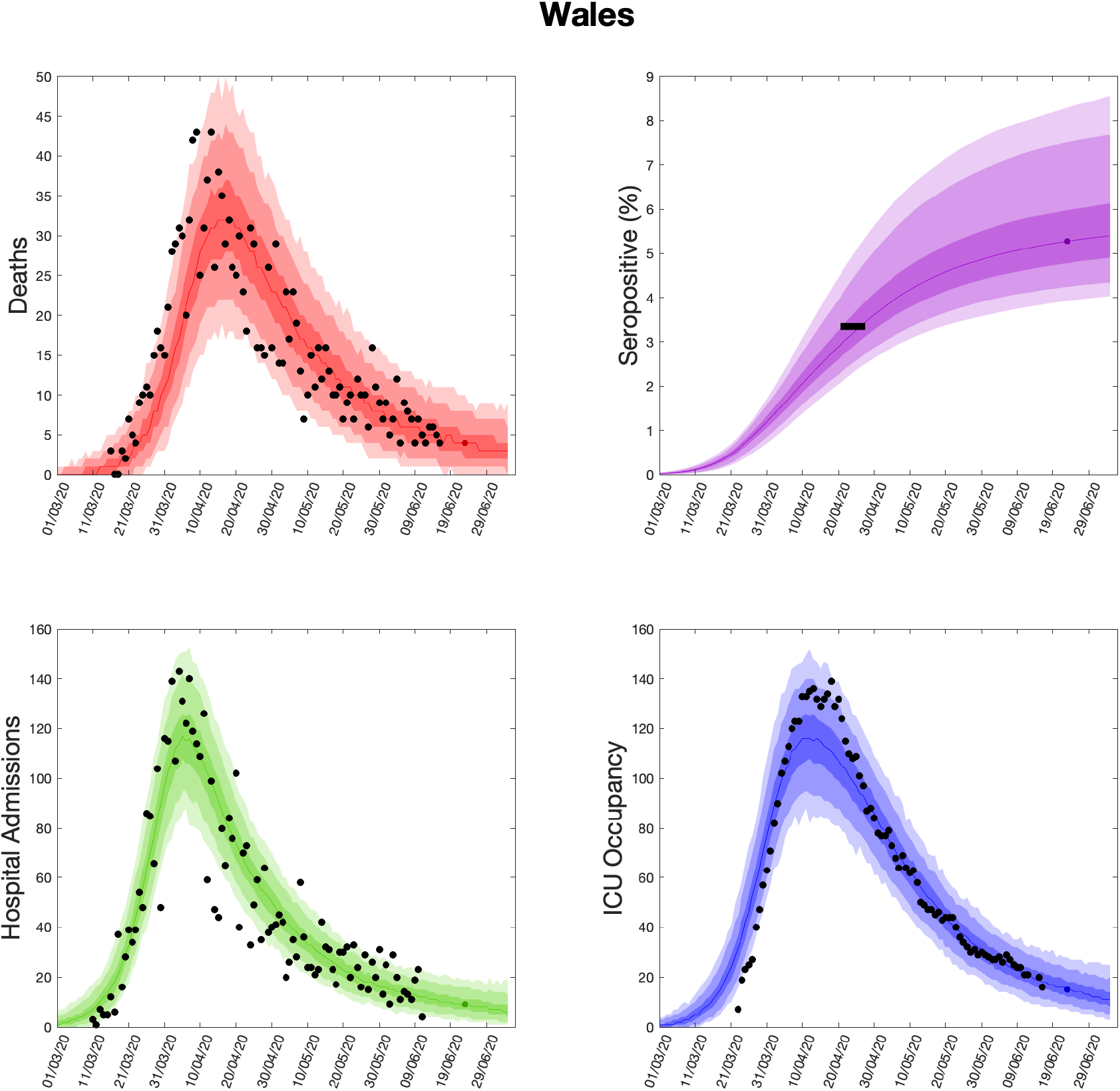
Health outcome predictions of the ODE from the beginning of the outbreak and 3 weeks into the future for Wales. **(Top left)** Daily deaths; **(top right)** seropositivity percentage; **(bottom left)** daily hospital admissions; **(bottom right)** ICU occupancy. In each panel: filled markers correspond to observed data, solid lines correspond to the mean outbreak over a sample of posterior parameters; shaded regions depict prediction intervals, with darker shading representing a narrower range of uncertainty (dark shading - 50%, moderate shading - 90%, light shading - 99%). The intervals represent our confidence in the fitted ODE model, and do not account for either stochastic dynamics nor the observational distribution about the deterministic predictions - which would generate far wider intervals. Predictions were produced using data up to 14th June 2020.

**Fig. S17:**
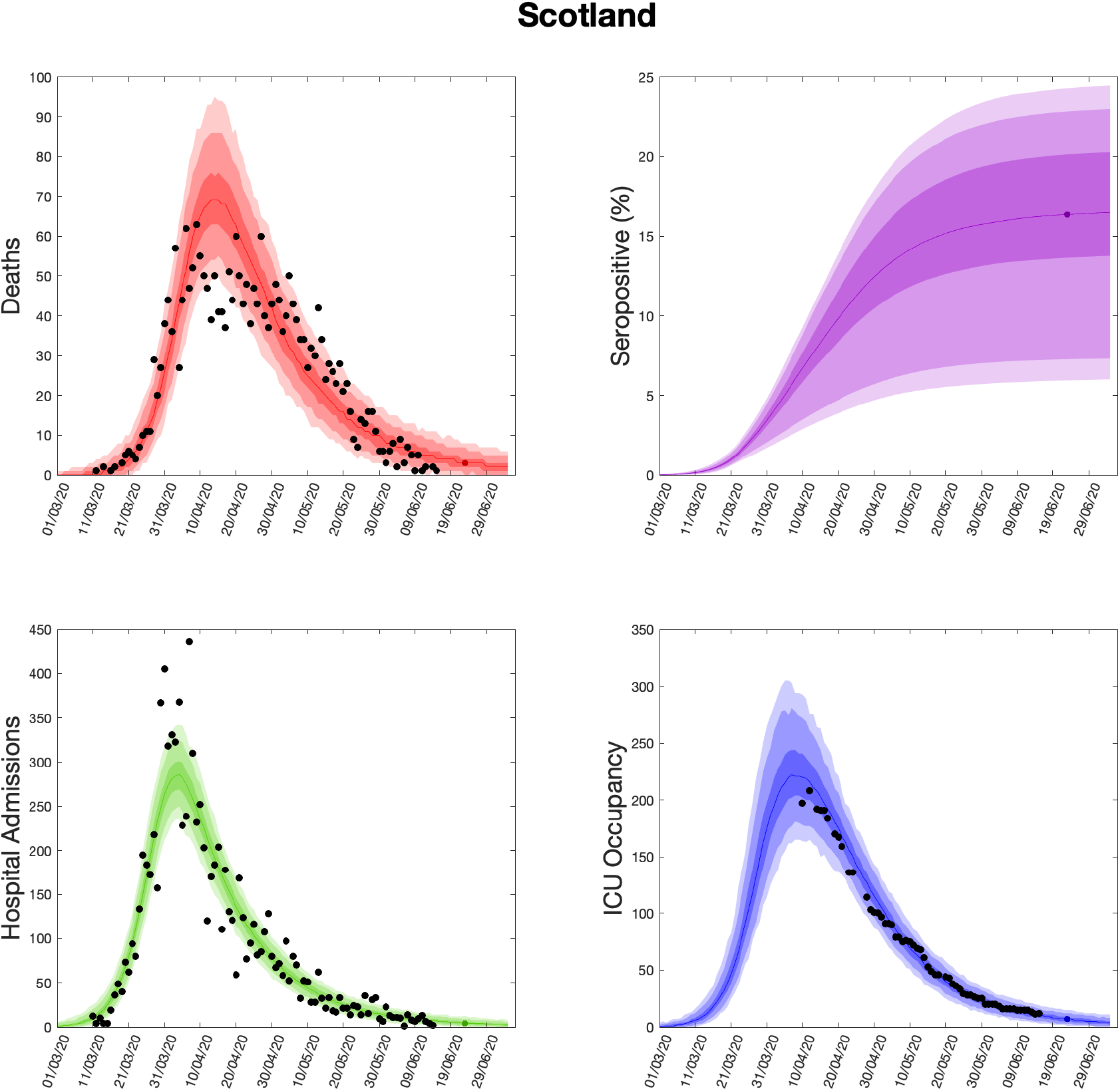
Health outcome predictions of the ODE from the beginning of the outbreak and 3 weeks into the future for Scotland. **(Top left)** Daily deaths; **(top right)** seropositivity percentage; **(bottom left)** daily hospital admissions; **(bottom right)** ICU occupancy. In each panel: filled markers correspond to observed data, solid lines correspond to the mean outbreak over a sample of posterior parameters; shaded regions depict prediction intervals, with darker shading representing a narrower range of uncertainty (dark shading - 50%, moderate shading - 90%, light shading - 99%). The intervals represent our confidence in the fitted ODE model, and do not account for either stochastic dynamics nor the observational distribution about the deterministic predictions - which would generate far wider intervals. Predictions were produced using data up to 14th June 2020.

**Fig. S18:**
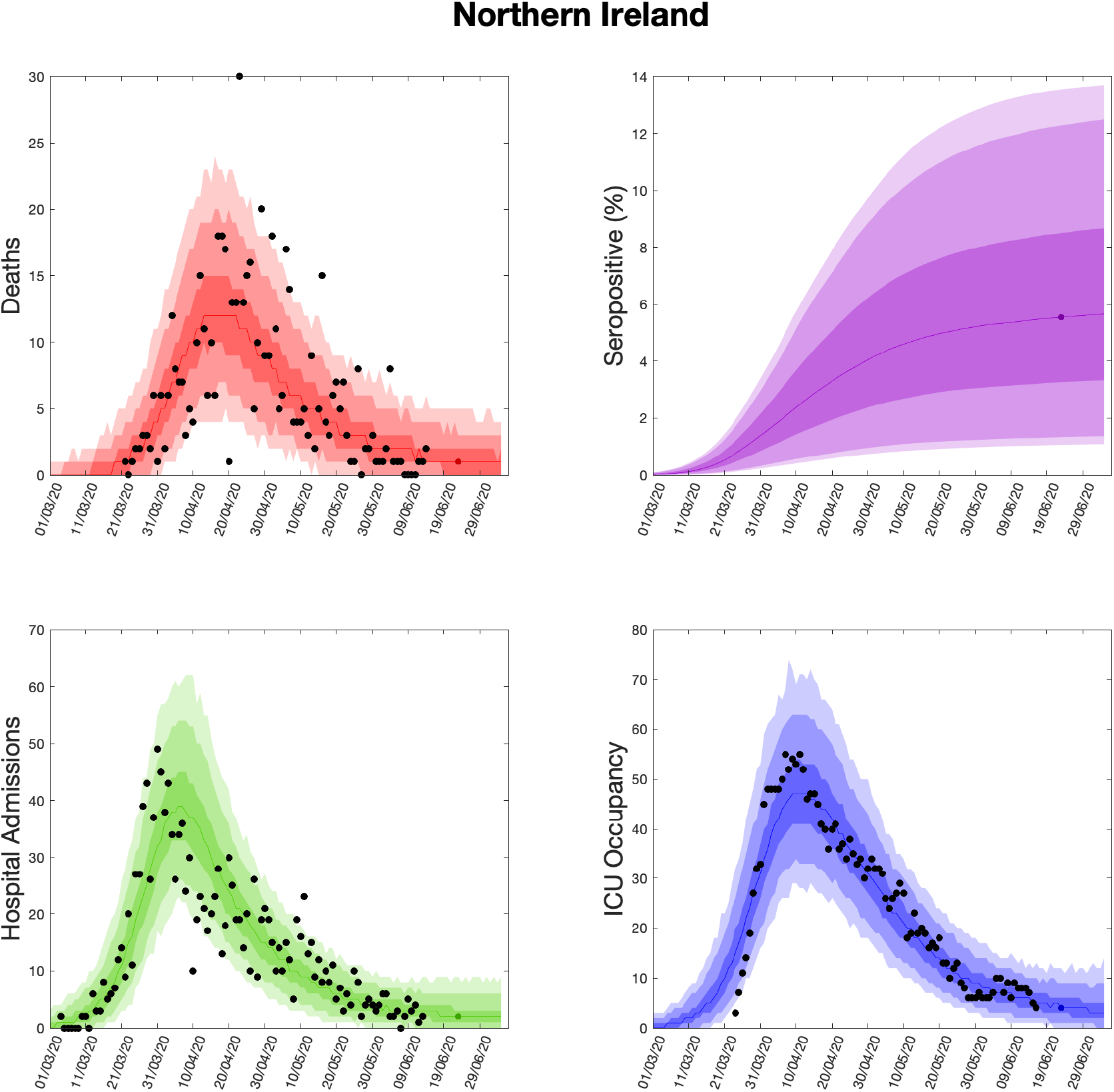
Health outcome predictions of the ODE from the beginning of the outbreak and 3 weeks into the future for Northern Ireland. **(Top left)** Daily deaths; **(top right)** seropositivity percentage; **(bottom left)** daily hospital admissions; **(bottom right)** ICU occupancy. In each panel: filled markers correspond to observed data, solid lines correspond to the mean outbreak over a sample of posterior parameters; shaded regions depict prediction intervals, with darker shading representing a narrower range of uncertainty (dark shading - 50%, moderate shading - 90%, light shading - 99%). The intervals represent our confidence in the fitted ODE model, and do not account for either stochastic dynamics nor the observational distribution about the deterministic predictions - which would generate far wider intervals. Predictions were produced using data up to 14th June 2020.

